# Identifying independent causal cell types for human diseases and risk variants

**DOI:** 10.1101/2024.05.17.24307556

**Authors:** Artem Kim, Zixuan Eleanor Zhang, Come Legros, Zeyun Lu, Adam J. de Smith, Jill E. Moore, Arun Durvasula, Nicholas Mancuso, Steven Gazal

## Abstract

The SNP-heritability of human diseases is extremely enriched in candidate regulatory elements (cREs) from disease-relevant cell types. Critical next steps are to understand whether these enrichments are driven by multiple causal cell types and whether individual variants impact disease risk via a single or multiple of cell types. Here, we propose CT-FM and CT-FM-SNP, 2 methods accounting for cREs shared across cell types to identify independent sets of causal cell types for a trait and its candidate causal variants, respectively. We applied CT-FM to 63 GWAS summary statistics (average *N* = 417K) using 924 cRE annotations, primarily from ENCODE4. CT-FM inferred 79 sets of causal cell types, with corresponding SNP-annotations explaining 39.0 ± 1.8% of trait SNP-heritability. It identified 14 traits with independent causal cell types, uncovering previously unexplored cellular mechanisms in height, schizophrenia and autoimmune diseases. We applied CT-FM-SNP to 39 UK Biobank traits and predicted high-confidence causal cell types for 3,091 candidate causal non-coding SNPs-trait pairs. Our results suggest that most SNPs affect a phenotype via a single set of cell types, whereas pleiotropic SNPs might target different cell types depending on the phenotype context. Altogether, CT-FM and CT-FM-SNP shed light on how genetic variants act collectively and individually at the cellular level to affect disease risk.

## Introduction

Understanding how genetic variants act collectively and individually at the cellular level to affect disease risk is critical to improve our understanding of disease biology ^1–3^. Previous studies have integrated cell-type-specific (CTS) functional annotations with genome-wide association studies (GWASs) to identify cell types presenting significant *association* with the disease or complex trait ^4–10^. However, most of these associated cell types are not truly causal (we define here a *causal cell type* as a cell type in which altered gene regulation affects disease risk), but only “tag” a disease causal cell type as a consequence of shared regulatory mechanisms across cell types and tissues ^11–15^. Thus, the number of cell types collectively targeted by disease variants, as well as how individual disease variants act at the cellular level (i.e., whether the variant affects disease risk by acting on a single causal cell type or on multiple causal cell types simultaneously) remain unclear. For example, we do not know which specific brain cell types targeted by GWAS variants contribute to body mass index (BMI) and obesity, what roles immune and adipose cell types may play ^16,17^, or how the FTO locus affects obesity risk through its actions in brain and adipose cell types ^18^. Altogether, these limitations hinder our understanding of how genetic variants confer disease risk at the cellular level.

Methods accounting for gene co-regulation in expression quantitative trait loci (eQTLs) datasets have already been proposed to infer causal tissues of human diseases ^14^ and their risk variants ^19^. However, current eQTL datasets have been generated on bulk tissues that do not capture CTS effects and often have limited overlap with GWAS results ^20–23^, thus limiting insights from these methods. Unlike eQTLs, candidate regulatory elements (cREs) are already available for thousands of cell types and conditions ^24–28^ and are extremely enriched in disease SNP-heritability (*h^2^*) when disease-relevant cell types have been assayed ^6,27–30^. However, we lack methods leveraging cREs and accounting for their shared structure across cell types to identify *independent* sets of causal cell types (i.e., cell types with limited cRE sharing).

Here, we propose CT-FM and CT-FM-SNP, methods that infer independent sets of causal cell types at the genome-wide and single variant level from CTS SNP-annotations, respectively. Both methods jointly analyze GWAS summary statistics with a set of CTS SNP-annotations. They output Bayesian posterior probabilities for each SNP-annotation to be causal for a trait (CT-FM) or a particular GWAS candidate variant (CT-FM-SNP) as well as independent causal sets (ICSs) reflecting the number of independent causal cell types. We applied both methods using 924 curated regulatory CTS SNP-annotations primarily from ENCODE4 (refs. ^24,31^) and single-cell ATAC-seq data from 30 fetal and adult tissues ^28^. We first validated and benchmarked CT-FM and CT-FM-SNP using simulations and GWAS of blood cell traits that biologically correspond to specific immune cell types. We applied CT-FM to a set of 63 independent GWASs (average *N* = 417K), identified expected causal cell types, and highlighted previously unexplored cellular mechanisms involved in height, schizophrenia (SCZ) and immune-related diseases. Finally, we applied CT-FM-SNP to 6,975 candidate causal {non-coding SNP, trait} pairs identified via SNP-fine-mapping within 39 UK Biobank traits ^32,33^. We found that most individual SNPs affected a phenotype via a single set of cell types and pleiotropic SNPs ^34^ might target different cell types depending on the phenotype context. Overall, CT-FM and CT-FM-SNP can be leveraged to infer independent sets of causal cell types and uncover new cellular mechanisms involved in disease risk.

## Results

### Overview of CT-FM and CT-FM-SNP

CT-FM (standing for Cell-Type-Fine-Mapping) and CT-FM-SNP are methods aiming to identify sets of independent causal cell types for a trait and trait-associated SNPs, respectively. They take as input a GWAS summary statistics, a matching LD reference panel, a set of CTS SNP-annotations, and (for CT-FM-SNP) a list of the trait’s candidate causal SNPs (**Fig. 1**).

**Figure 1.**
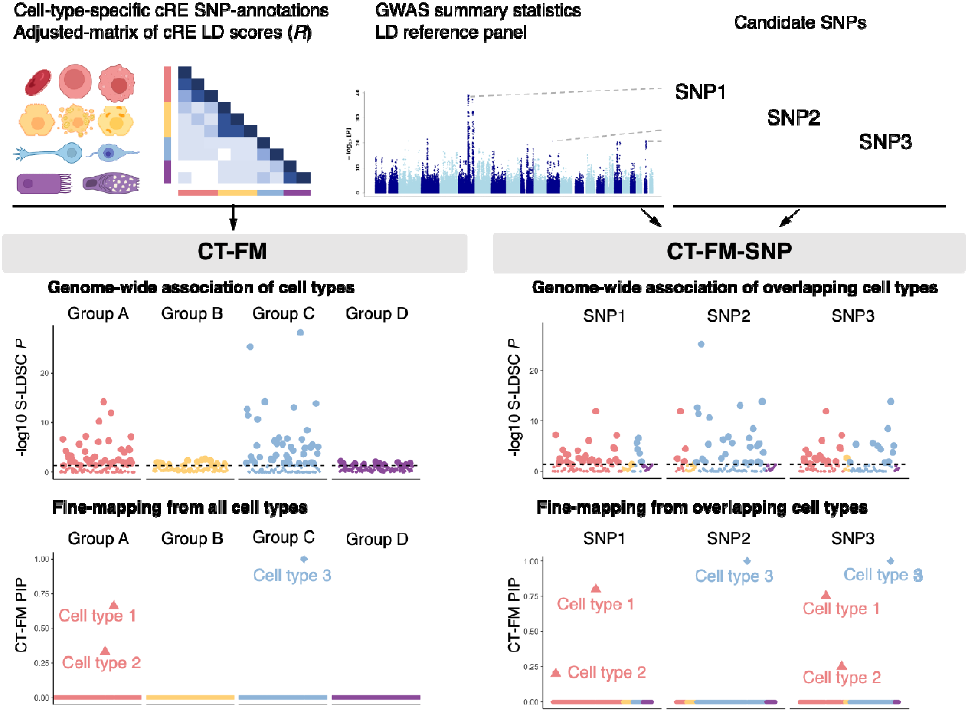
Overview of the CT-FM framework. CT-FM and CT-FM-SNP are 2 methods that identify sets of independent causal cell types of a trait and its candidate causal SNPs, respectively. They take as inputs a set of cell-type-specific (CTS) candidate regulatory elements (cREs) SNP-annotations, GWAS summary statistics with a matching LD reference panel, and (for CT-FM-SNP only) a list of a trait’s candidate causal SNPs. First, CT-FM estimates the significance of the marginal effect on SNP-heritability of each CTS SNP-annotation by applying stratified LD score regression (S-LDSC) ^29,35,36^. Then, CT-FM infers the causal cell types and outputs posterior inclusion probability (PIP) and independent causal sets (ICSs) by leveraging S-LDSC Z-scores and an adjusted-matrix of cRE linkage disequilibrium (LD) scores of CTS SNP-annotations. In our toy example, CT-FM reduces the number of SNP-annotations significantly associated with the trait to 2 ICSs (triangle and diamond), each corresponding to cell types of a distinct biological group (A and C). CT-FM-SNP leverages the same workflow as CT-FM, in the difference that it restricts the inference procedure to CTS SNP-annotations that overlap the candidate SNP. In our toy example, CT-FM-SNP infers the causal cell types of 3 GWAS candidate SNPs and assigns the first candidate SNP to cell types from the biological group A, the second candidate SNP to a cell type of the biological group C, and the third candidate SNP to cell types of the biological groups A and C. The dashed horizontal line represents the S-LDSC significance threshold.

CT-FM aims to identify independent sets of CTS SNP-annotations that are the most likely to explain the *h^2^* observed across a large set of CTS SNP-annotations. It models the vector of per-normalized-genotype effect sizes *β* as a mean 0 vector with variance *Var*(*β _j_*) = ∑_*b*∈ *B*_a_b_ (*j*) *τ _b_*+ ∑_*C*∈*C*_a_C_ (*j*) *τ_C_*, where *B* represents a set of background (non-CTS) SNP-annotations (including coding, enhancer, and promoter SNP-annotations from the baseline model ^4,6^), *a_b_* (*j*) is the indicator variable of variant *j* for SNP-annotation *b*, _b_is the contribution of *b* to *Var*(*β_j_*), and *C* represents the set of CTS SNP-annotations. CT-FM assumes that at most, *L* SNP-annotations contribute positively to *Var*(*β_j_*). It models the vector *τ* of *τ*_*C*∈**C**_as 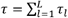 as the sum of *L* single-effects *τ_l_* = *γ_l_* · *b_l_* where *γ_l_* is a *C* × 1 binary vector indicating which CTS SNP-annotation is causal for the *l^th^*effect, and *b_l_* is a scalar quantifying the contribution of the causal *l^th^*effect to *Var*(*β_j_*). CT-FM applies the following procedure to infer *L* and the most likely causal cell types. First, for each CTS SNP-annotation *c*, it applies stratified LD score regression (S-LDSC) ^29,35,36^ with the *B* background SNP-annotations to estimate their marginal effect 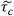. Second, it assigns Bayesian posterior probabilities to SNP-annotations from the vector of marginal 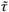 Z-scores using an extension of the Sum of Single Effects (SuSiE) model ^37,38^ on a *adjusted-matrix of cRE LD scores R* defined as the correlation of the LD scores of the CTS SNP-annotations adjusted on the *B* background SNP-annotations. CT-FM outputs posterior inclusion probabilities (PIPs) for every CTS SNP-annotation, which correspond to probabilities of each corresponding cell type to be causal for the trait. It also regroups putative causal correlated SNP-annotations (i.e., cell types presenting highly shared cREs) within *L* independent causal sets (ICSs), thus estimating the number of independent sets of cell types while accounting for uncertainty in cell type prioritization. Here, CT-FM defined a *candidate* causal cell type when a corresponding SNP-annotation was found in an ICS and a *highly-confident* causal cell type when a corresponding SNP-annotation had a PIP ≥ 0.5; when no highly-confident causal cell type was detected but multiple CTS SNP-annotations from the same cell type were detected in the same ICS (for example, several SNP-annotations corresponding to B cells stimulated in different context), we reported a combined PIP (cPIP; see **Methods**) for the cell type (i.e., B cells). We note that although CT-FM generalizes the SuSiE SNP-fine-mapping model, the methods differ in several key ways (see **Discussion** and **Methods**).

CT-FM-SNP aims to identify the most likely causal cell types of a trait candidate causal SNP by leveraging both polygenic enrichment within overlapping SNP-annotations and co-regulation within cREs. It applies the same workflow as CT-FM but differs in that it restricts the inference procedure to marginal 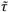 Z-scores of CTS SNP-annotations that overlap the candidate SNP. Similar to CT-FM, CT-FM-SNP defined candidate causal cell types for a trait candidate SNP when their corresponding SNP-annotations were found in an ICS and highly-confident causal cell types for a trait candidate SNP when their corresponding SNP-annotations had a PIP ≥ 0.5. Although the inference of multiple ICSs by CT-FM suggests that independent cell types are causal for the disease (as it relies on polygenicity), the inference of multiple ICSs by CT-FM-SNP suggests that the candidate SNP putatively targets multiple independent causal cell types but does not allow for concluding whether the candidate SNP affects disease risk by disrupting gene regulation within a single cell type or multiple cell types.

We applied CT-FM and CT-FM-SNP with a total of 924 CTS SNP-annotations (capturing, on average, 1.1% of common SNPs) derived from cREs of human cell types and human-derived cell lines (see **Fig. 1** and **Methods**), classified into 9 biological groups for visualization (**Supplementary Fig. 1**). CTS SNP-annotations were cREs from ENCODE4 (650), CATlas single-cell ATAC-seq data ^28^ (222) and the ABC method ^26^ (52) (**Supplementary Table 1**). CT-FM was successfully applied (i.e., detected at least one ICS) to 63 independent and well-powered GWASs prioritizing diseases over quantitative traits (average *N* = 417K; see **Methods** and **Supplementary Table 2**). We investigated the contribution of each ICS to trait *h*^2^ by creating SNP-annotations in which we merged its constituent SNP-annotations. CT-FM-SNP was applied to 6,975 {non-coding variant, trait} pairs obtained via SNP-fine-mapping on 39 UK Biobank traits ^32,33^. To further predict causal {non-coding SNP, cell type, gene, trait} quadruplets, we leveraged predictions from both CT-FM-SNP and SNP-gene links from our cS2G strategy ^39^.

**Table 1.**
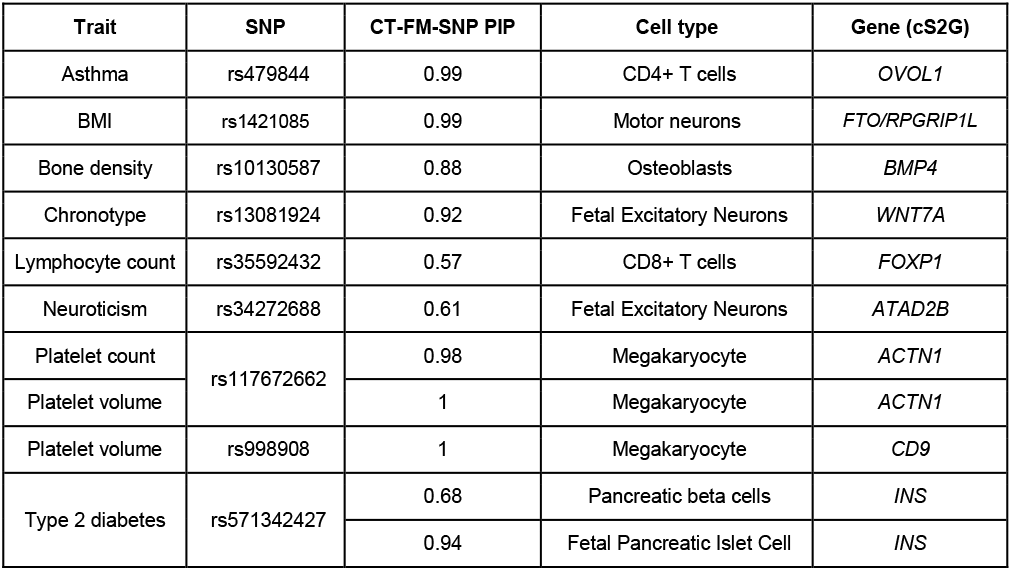
Notable {non-coding SNP, cell type, gene, trait} quadruplets. Results for rs1421085, rs10130587, rs34272688 and rs571342427 are discussed in the main text. Results for other candidate SNPs are discussed in the **Supplementary Note**.

| Trait | SNP | CT-FM-SNP PIP | Cell type | Gene (cS2G) |
| --- | --- | --- | --- | --- |
| Asthma | rs479844 | 0.99 | CD4+ T cells | *OVOL1* |
| BMI | rs1421085 | 0.99 | Motor neurons | *FTO/RPGRIP1L* |
| Bone density | rs10130587 | 0.88 | Osteoblasts | *BMP4* |
| Chronotype | rs13081924 | 0.92 | Fetal Excitatory Neurons | *WNT7A* |
| Lymphocyte count | rs35592432 | 0.57 | CD8+ T cells | *FOXP1* |
| Neuroticism | rs34272688 | 0.61 | Fetal Excitatory Neurons | *ATAD2B* |
| Platelet count | rs117672662 | 0.98 | Megakaryocyte | *ACTN1* |
| Platelet volume |  | 1 | Megakaryocyte | *ACTN1* |
| Platelet volume | rs998908 | 1 | Megakaryocyte | *CD9* |
| Type 2 diabetes | rs571342427 | 0.68 | Pancreatic beta cells | *INS* |
|  |  | 0.94 | Fetal Pancreatic Islet Cell | *INS* |

Further details are provided in **Methods**. We have released open-source software implementing our framework (see **Code availability**) and all CTS SNP-annotations and GWAS summary statistics analyzed are publicly available (see **Data availability**).

### Validating and benchmarking CT-FM and CT-FM-SNP using simulations

We performed extensive simulations to assess CT-FM and CT-FM-SNP power to identify causal cell types. Specifically, we simulated 500 GWAS summary statistics using LD patterns from the UK Biobank and effect sizes depending on per-SNP *h*^2^ estimated by S-LDSC on a height GWAS ^40^. We considered a sample size of *N* = 350K (same order of magnitude as GWASs analyzed by CT-FM and CT-FM-SNP) and *h*^2^ = 0.5 because these 2 parameters provided a maximum S-LDSC Z-score distribution similar to the one observed in the GWASs further analyzed by CT-FM and CT-FM-SNP (**Supplementary Fig. 2**). We considered primarily 2 SNP-annotations corresponding to osteoblasts and/or fibroblasts as causal (identified by CT-FM as causal for height, see *Novel findings from traits with independent sets of causal cell types* section). Methods were evaluated using precision (the proportion of SNP-annotations inferred as causal that are truly causal for a trait/SNP), sensitivity (the proportion of causal SNP-annotations inferred as causal for a trait/SNP), and F1 scores. Further details are provided in **Methods**.

We first evaluated CT-FM power to identify causal cell types of a trait in simulations in which we varied the number of causal SNP-annotations from 1 to 3 (**Fig. 2** and **Supplementary Table 3**). We primarily evaluated 4 approaches: CT-FM-candidate (reporting SNP-annotations in CT-FM ICSs), CT-FM-high-confidence (reporting SNP-annotations with CT-FM PIP > 0.5), S-LDSC-candidate (reporting SNP-annotations with an S-LDSC false discovery rate (FDR) *P* < 0.05), and S-LDSC-topFDR (reporting the SNP-annotation with the most significant S-LDSC FDR *P*); more methods based on S-LDSC FDR *P* were explored in **Supplementary Fig. 3**. Overall, the sensitivity of all methods decreased with increasing number of causal cell types, but precision remained stable. CT-FM-high-confidence maximized precision (≥0.79), achieved a high F1 score in simulations considering one causal cell type (0.85, similar to the highest F1 score of 0.87 obtained by S-LDSC-topFDR), and maximized F1 scores (≥0.53) in simulations considering multiple causal cell types, making CT-FM-high-confidence the strategy of choice to infer causal cell types. CT-FM-candidate provided lower sensitivities than S-LDSC-candidate but higher F1 scores, making it the strategy of choice to nominate candidate cell types while maximizing sensitivity. Additionally, CT-FM provided high PIP values for the causal annotation (average 0.77, 0.54, and 0.42 in simulations with 1, 2 and 3 causal annotations, respectively) and detected multiple ICSs in simulations considering multiple causal cell types (33% and 71% of simulations with 2 and 3 causal annotations, respectively). In simulations with diverse single causal SNP-annotations, precision and sensitivity decreased when the causal SNP-annotation had a high *R* value with other analyzed SNP-annotations (**Supplementary Figs. 4**-**5**); in simulations in which CT-FM did not provide the highest PIP value for the causal SNP-annotation, the inferred causal annotation was still highly correlated with the causal one (mean *R* ≥ 0.48, indicating that when CT-FM fails to identify the causal cell type, it still pinpoints a highly correlated cell type (**Supplementary Fig. 6**).

**Figure 2.**
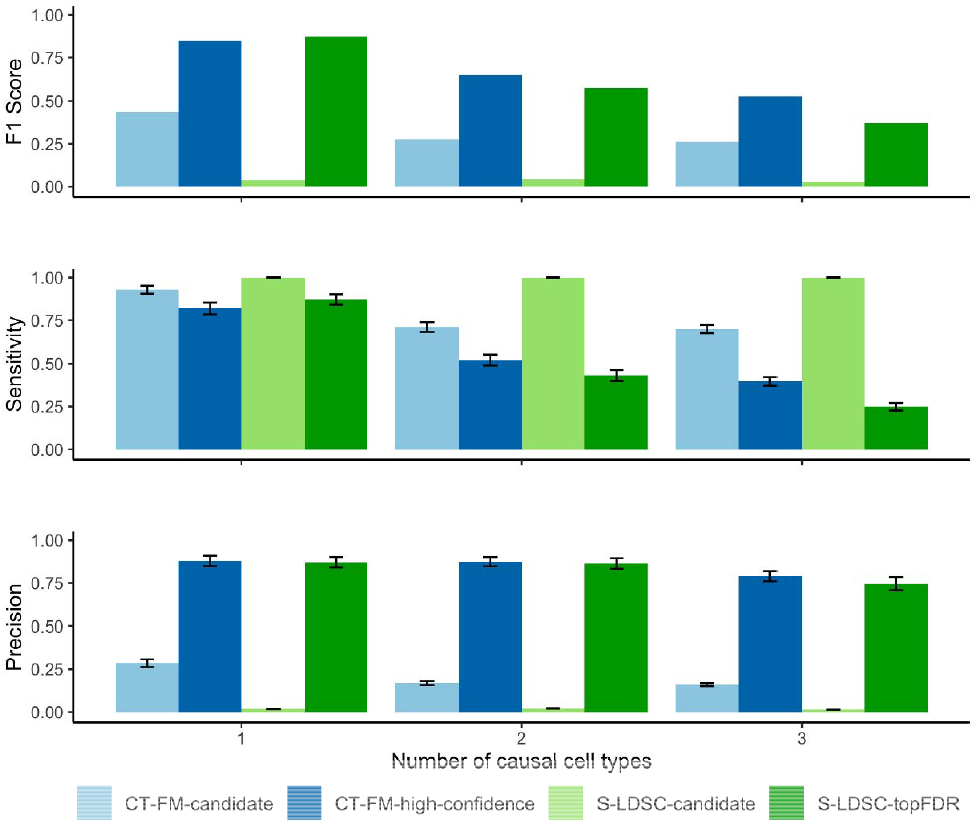
Simulations to assess precision and sensitivity of methods inferring causal cell types. We report the precision, sensitivity and F1 score in simulations with different numbers of causal SNP-annotations. Error bars represent 95% confidence intervals. Numerical results are reported in **Supplementary Table 3**.

We performed 3 additional sets of simulations considering 2 causal SNP-annotations. First, we replicated conclusions from above by performing simulations in which we let vary *N* (**Supplementary Fig. 7**) and *h*^2^ (**Supplementary Fig. 8**); as expected, CT-FM performance improved when increasing these 2 parameters. Second, assuming that the causal cell type has in practice not been assayed, we performed simulations in which we added noise in the causal SNP-annotation; CT-FM results were robust to a high level of noise (20%) in the causal SNP-annotations (**Supplementary Fig. 9**). Finally, we replicated our conclusions when varying the *h*^2^ explained by the 2 causal SNP-annotations (**Supplementary Fig. 10**).

We next evaluated CT-FM-SNP power to identify causal cell types of candidate SNPs for a trait (**Fig. 3** and **Supplementary Table 4**). We evaluated 4 approaches: CT-FM-SNP-candidate, CT-FM-SNP-high-confidence (reporting similar SNP-annotations as their corresponding CT-FM approaches), S-LDSC-SNP-candidate (reporting SNP-annotations overlapping the candidate SNP with S-LDSC FDR *P* < 0.05), and S-LDSC-SNP-topFDR (reporting the SNP-annotation overlapping the candidate SNP with the smallest S-LDSC FDR *P* < 0.05). We evaluated type I error by using 500 candidate SNPs that did not overlap the causal SNP-annotation(s) and evaluated precision, sensitivity and F1 scores using 500 candidate SNPs that overlapped the causal SNP-annotation(s); candidate SNPs were sampled according to their expected per-SNP *h*^2^ and their overlap with causal SNP-annotation(s). First, we performed simulations in which we considered osteoblasts as the single causal cell type. CT-FM-SNP-high-confidence had significantly lower type I error (0.075) than S-LDSC-related methods (≥0.29) (**Fig. 3a**) while maintaining high precision (0.90) and high F1 score (0.87) (**Fig. 3b**). Second, we performed simulations in which we considered osteoblasts and fibroblasts as the 2 causal cell types. CT-FM-SNP-high-confidence still had significantly lower type I error (0.11) than S-LDSC-related methods (≥0.31) (**Fig. 3c**) while maintaining moderate precision (≥0.56) and F1 score (≥0.53) (**Fig. 3d** and **Supplementary Fig. 11**). Finally, we focused on the 263/500 simulations in which CT-FM identified a single ICS corresponding to fibroblasts and failed to identify osteoblasts as the second causal cell type and investigated whether CT-FM-SNP could identify this second causal cell type for candidate SNPs overlapping the osteoblast (but not the fibroblast) SNP-annotation (**Supplementary Fig. 12**). CT-FM-SNP was able to assign >10% (resp. 25% and 50%) of the candidate SNPs to the osteoblast annotation with high confidence in 78% (resp. 38% and 27%) of the cases. Higher performance was observed when interverting the osteoblast and fibroblast annotations (i.e., 91%, 91% and 79%, respectively; **Supplementary Fig. 13**). Overall, these results highlight CT-FM-SNP-high-confidence as the strategy of choice to infer causal cell types of candidate SNPs and that CT-FM-SNP can nominate causal cell types that have been missed by CT-FM.

**Figure 3.**
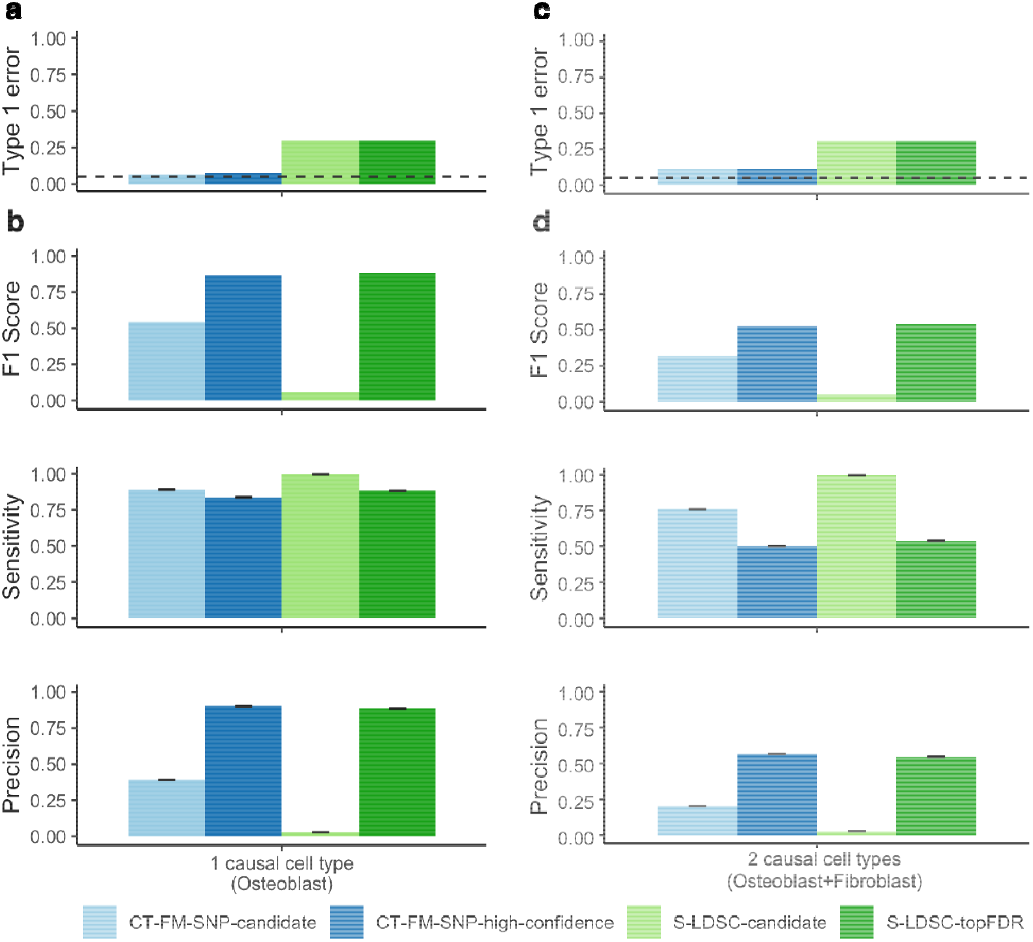
Simulations to assess type I error, precision and sensitivity of methods inferring causal cell types of candidate SNPs. We report the type I error (**a,c**), the precision, sensitivity and F1 score (**b,d**) in simulations in which we considered only osteoblasts (**a,b**) and osteoblasts and fibroblasts (**c,d**) as causal cell types. We report results for candidate SNPs overlapping the osteoblast SNP-annotation in (**d**); results for candidate SNPs overlapping the fibroblast SNP-annotation and overlapping both osteoblast and fibroblast SNP-annotations are reported in **Supplementary Fig. 11**. Error bars represent 95% confidence intervals. Numerical results are reported in **Supplementary Table 4**.

Altogether, these simulations demonstrate that CT-FM and CT-FM-SNP are powerful and precise methods to infer the causal CTS SNP-annotations from well-powered GWAS. Although in the following, we primarily focus our results on high-confidence cell types (because of high precision), we also report CT-FM candidate cell types in Figures and the cell type with the highest PIP, if necessary.

### Validating and benchmarking CT-FM and CT-FM-SNP on blood traits

As proof of principle on real GWAS datasets, we further applied CT-FM and CT-FM-SNP to 5 blood cell traits that biologically correspond to specific immune cell types ^5,41,42^ (**Fig. 4**).

We applied CT-FM to the 5 traits using GWAS summary statistics from the UK Biobank ^40^ (average *N* = 444K) and identified expected causal cell types (**Fig. 4a** and **Supplementary Table 5**). Specifically, CT-FM identified one ICS per trait (average of 4.4 CTS SNP-annotations per trait), all highlighting a high-confidence causal cell type (PIP ≥ 0.5), such as T cells for lymphocyte count (PIP = 0.59 for CD8+ T cells) and monocytes for monocyte count (PIP = 0.72). CT-FM greatly reduced the number of candidate causal cell types as compared with the initial association signal inferred by S-LDSC (**Fig. 4b** and **Supplementary Table 6**). We performed additional CT-FM benchmarking analyses investigating the number of ICSs (**Supplementary Fig. 14**), SuSiE parameters *L* and sample size *n* (**Supplementary Table 5**), and the use of SuSiE-inf ^43^ (**Supplementary Table 7**).

We next applied CT-FM-SNP on 1,564 candidate causal {non-coding variant, trait} pairs of the 5 blood traits (obtained by SNP-fine-mapping in the UK Biobank ^32,33^ and by selecting SNPs with a SNP-PIP > 0.5). We identified at least one ICS for nearly half of the candidate SNPs (48%, 758 of 1,564; 829 ICSs in total) and detected a high-confidence causal cell type for 90% of these ICSs (748 ICSs with PIP > 0.5 among the 829 ICSs), highlighting a high success rate of CT-FM-SNP (**Fig. 4c** and **Supplementary Table 8**). Among the CT-FM-SNP ICSs with a high-confidence causal cell type, 76% (**571** of **748**) corresponded to candidate cell types previously identified by CT-FM, thus demonstrating high consistency of causal cell types inferred by CT-FM and CT-FM-SNP (**Fig. 4d, Supplementary Table 9**). For the platelet count trait, we identified 66 ICSs with a high-confidence causal cell type inferred by CT-FM-SNP but not found in CT-FM ICSs (**Fig. 4d**); approximately half of these high-confidence causal cell types (53%, **35** of **66**) corresponded to the megakaryocyte cell type (**Supplementary Table 10**, see also **Supplementary Fig. 14**), thus confirming (from simulations) that in cases in which CT-FM fails to detect a causal cell type, this cell type can be captured by CT-FM-SNP. Similar conclusions for the red blood cell (RBC) volume trait are discussed in **Supplementary Table 10**. Similar patterns were observed when restricting analyses to SNPs with SNP-PIP > 0.95 (282 SNPs in total), which demonstrates that our conclusions are robust to the imperfect selection of candidate causal SNPs (**Supplementary Fig. 15**).

We functionally validated the pairs identified by CT-FM-SNP for lymphocyte and monocyte count candidate SNPs using single-cell *cis*-eQTLs from OneK1K ^44,45^ (**Fig. 4e-f** and **Supplementary Table 11**). Specifically, lymphocyte count candidate SNPs linked to lymphocyte cell types by CT-FM-SNP exhibited a higher proportion of lymphocyte single-cell *cis*-eQTLs as compared to a set of control SNPs (see **Methods**) and as well as a set of lymphocyte count candidate SNPs not assigned to lymphocyte cell types by CT-FM-SNP (**Fig. 4e**). Similar conclusions were observed for monocyte count candidate SNPs and monocyte single-cell *cis*-eQTLs (**Fig. 4f**).

**Figure 4.**
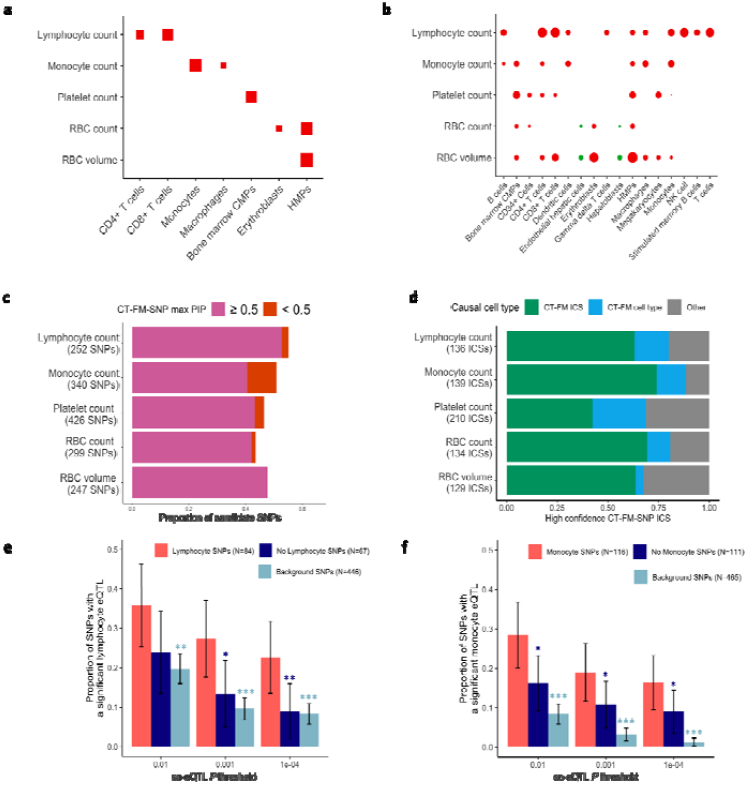
Benchmarking CT-FM and CT-FM-SNP on 5 blood cell traits. **(a)** We report CT-FM candidate causal cell types. Dot sizes are proportional to CT-FM PIP. Numerical results are reported in **Supplementary Table 5. (b)** We report cell types identified by CT-FM as significantly associated with each trait. Dot sizes are proportional to S-LDSC -log_10_ FDR *P* values, and dot colors represent biological groups (red for blood/immune and green for digestive). Only CTS SNP-annotations with S-LDSC FDR *P* value < 0.05 are represented. Numerical results are reported in **Supplementary Table 6. (c)** We report the proportion of candidate causal SNPs that were linked to at least one causal cell type by CT-FM-SNP. Results for all candidate variants are reported in **Supplementary Table 8. (d)** We report the proportion of high-confidence {non-coding SNP, cell type, trait} triplets inferred by CT-FM-SNP for which the cell type is consistent with CT-FM results. We highlight triplets for which the causal CTS SNP-annotation was also found in CT-FM ICSs (green), triplets for which the causal CTS SNP-annotation was not found in CT-FM ICSs but corresponds to the same cell type (blue), and triplets for which the causal CTS SNP-annotation was not found in CT-FM ICSs (grey). Numerical results are reported in **Supplementary Table 9. (e)** We report the fraction of SNPs with a lymphocyte single-cell *cis*-eQTL for lymphocyte count candidate SNPs assigned to lymphocyte cell type by CT-FM-SNP (red), candidate SNPs not assigned to lymphocyte cell type (dark blue), and a set of background SNPs (light blue) (* *P* < 0.05; ** *P* < 0.01; *** *P* < 0.001, one-tailed). **(f)** Similar to **(e)** with monocyte single-cell *cis*-eQTLs and monocyte count candidate SNPs. Numerical results are reported in **Supplementary Table 11**. RBC: red blood cell; CMPs: common myeloid progenitor cells; HMPs: hematopoietic multipotent progenitors; NK: natural killer.

Finally, we quantified the benefits of leveraging CT-FM and CT-FM-SNP compared to methods leveraging gene expression datasets. First, we used S-LDSC to compare the *h*^2^ explained by SNP-annotations constituted by CT-FM ICSs with the *h*^2^ explained by SNP-annotations based on fine-mapped eQTLs from GTEx ^21,46^ and OneK1K ^44,45^. CT-FM ICSs explained at least 4.3x more *h*^2^ than fine-mapped eQTLs (48.2 ± 5.7% vs. 8.5 ± 0.9% and 11.1 ± 1.0% for GTEx and OneK1K, respectively; **Supplementary Table 12**). Second, we compared high-confidence CT-FM-SNP and cS2G results to those reported by TGFM ^19^ using GTEx (TGFM-GTEx) or single-cell data from peripheral blood mononuclear cells ^47^ (TGFM-sc) datasets. The number of unique {cell type,gene} pairs identified by CT-FM-SNP+cS2G was 3.7x greater than the number of {tissue,gene} pairs reported by TGFM-GTEx and 65.9x greater than the number of {cell type,gene} pairs reported by TGFM-sc (**Supplementary Table 13**). For example, CT-FM-SNP+cS2G identified 122 pairs for the lymphocyte count GWAS (including 76 in CD8+ T cells), whereas TGFM-GTEx reported 35 pairs (including 20 in whole blood) and TGFM-sc reported 1 pair (CD4+ T cells).

Altogether, our results demonstrate that CT-FM and CT-FM-SNP were able to capture known cellular mechanisms of 5 blood cell traits. Of note, CT-FM-SNP and cS2G pinpointed {SNP-gene-cell-type} triplets consistent with single-cell cis-eQTLs from OneK1K and nominated more precise causal mechanisms than state-of-the-art methods leveraging gene expression datasets, even when large-scale single-cell datasets in relevant cell types are available.

### Applying CT-FM to 63 GWASs

We successfully applied CT-FM to a total of 63 independent well-powered GWASs (average *N* = 419K, see **Methods**) and report our main findings in **Fig. 5**. CT-FM identified 79 ICSs (average of 7.25 CTS SNP-annotations per ICS) and 56 high-confidence causal cell types (**Fig. 5a** and **Supplementary Table 14**); for the 23 ICSs without a high-confidence causal cell type, we identified 7 cell types with cPIP > 0.5 (**Supplementary Table 15**). CT-FM highlighted independent sets of causal cell types (i.e., at least 2 ICSs) for 14 complex traits (**Fig. 5a**; new insights from these traits are discussed in the next section). CT-FM identified an average of 9.1 candidate CTS SNP-annotations per trait, representing a ∼20x decrease from candidate SNP-annotations identified by S-LDSC (FDR *P* < 0.05) (**Supplementary Fig. 16** and **Supplementary Table 16**); as an illustration, CT-FM significantly decreased the initial association signal of S-LDSC for SCZ across 8 different biological groups (484 SNP-annotations with FDR *P* < 0.05) to 3 ICSs across brain and immune cell types (**Fig. 5b** and **Supplementary Table 17**).

**Figure 5.**
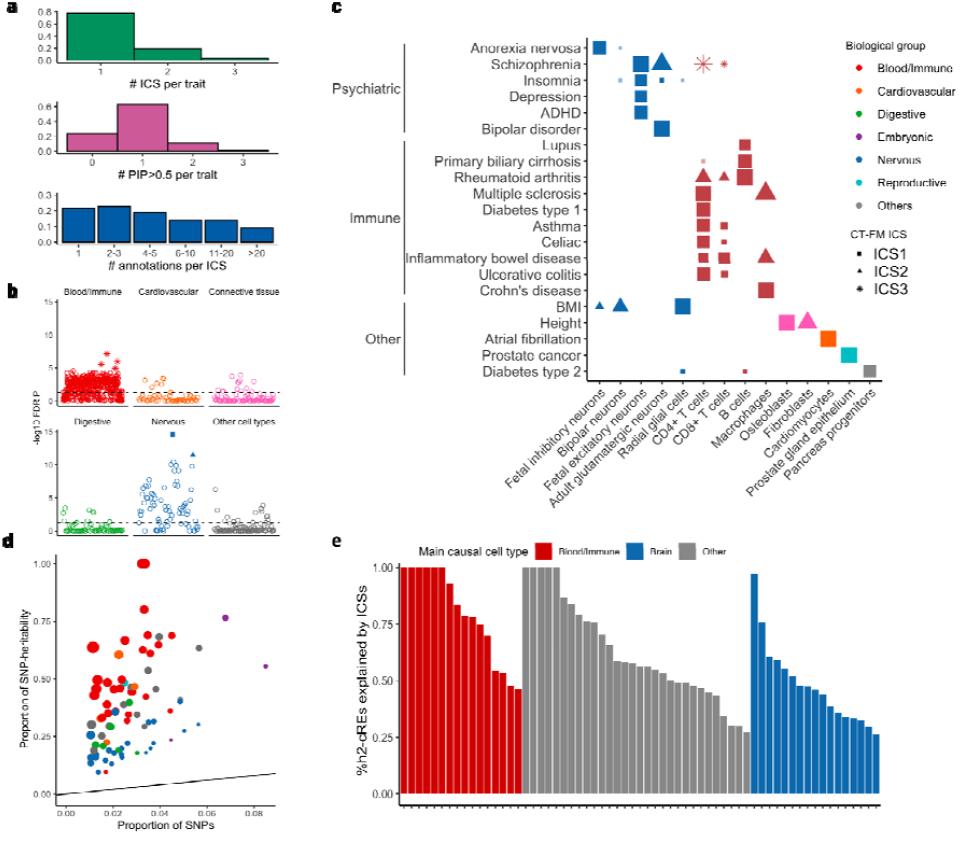
Application of CT-FM to 63 GWAS summary statistics. **(a)** We report the number of ICSs per trait, highly-confident causal cell types (PIP > 0.5) per trait, and CTS SNP-annotations per ICS. Causal cell types with PIP < 0.5 but cPIP > 0.5 were not reported. Numerical results are reported in **Supplementary Table 14. (b)** CT-FM and S-LDSC results for schizophrenia. For each CTS SNP-annotation, we report S-LDSC -log_10_ FDR *P* values on the axis, and CT-FM ICSs by different shapes (square for fetal excitatory neurons, triangle for adult glutamatergic neurons, and asterisk for immune cell types; SNP-annotations not in ICS are represented with open circle). The dashed horizontal line represents the S-LDSC FDR significance threshold. S-LDSC results for the 924 CTS SNP-annotations are reported in **Supplementary Table 17. (c)** We report notable candidate causal cell types inferred by CT-FM for different complex traits. Dot sizes are proportional to CT-FM PIP. CT-FM results for the 63 GWASs are reported in **Supplementary Tables 14-15. (d)** We report the proportion of *h*^2^ explained by each CT-FM ICSs. Dot sizes are proportional to *h*^2^ enrichment and dot colors represent biological groups (same as **c**). Numerical results are reported in **Supplementary Table 18. (e)** We report the proportion of *h*^2^-cREs explained by CT-FM ICSs for each trait. Numerical results are reported in **Supplementary Table 19**. Proportions of *h*^2^ and *h*^2^-cREs > 1 were rounded in (**e,f**) for visualization purposes; we note that values > 1 are outside the biologically plausible 0-1 range, but allowing point estimates outside the biologically plausible 0– 1 range is necessary to ensure unbiasedness. ADHD: attention-deficit/hyperactivity disorder; BMI: body mass index.

CT-FM identified high-confidence causal cell types consistent with known disease biology (**Fig. 5c**). For psychiatric diseases, CT-FM identified excitatory neurons for SCZ ^48,49^ (PIP = 0.98) and bipolar disorder ^50^ (PIP = 0.99), excitatory neurons for insomnia ^51^ (cPIP = 0.47), depression ^52^ (PIP = 0.45), and attention-deficit/hyperactivity disorder ^53^ (PIP = 0.58), and inhibitory neurons for anorexia ^54^ (PIP = 0.64). It also inferred B cells and/or T cells for multiple immune-related diseases, cardiomyocytes for atrial fibrillation ^55^ (PIP = 0.99), pancreatic cells for type 2 diabetes ^56^ (PIP = 0.51) and prostate epithelial cells for prostate cancer ^57^ (PIP = 0.99).

We created SNP-annotations corresponding to each ICS (average of 2.7% of common SNPs) and found that they explained a high fraction of *h*^2^ (34.5 ± 1.5% of *h*^2^ explained by each ICS SNP-annotation and 39.0 ± 1.8% of *h*^2^ explained by the union of ICS SNP-annotations; **Fig. 5d** and **Supplementary Tables 18-19**). Despite these high enrichments, CT-FM identified a relatively small number of ICSs per trait (1.25; 49 of 63 traits with a single ICS), which suggests missing causal cellular mechanisms (consistent with simulations). We thus quantified CT-FM success to create ICSs capturing most of causal cell types using h2 analyses. Specifically, we computed, for each trait, the *h*^2^ explained by the largest available cRE datasets ^24–28,46^ (*h*^2^-cREs; 38.9% of common SNPs accounting for 60.6 ± 1.5% of *h*^2^ across 63 traits; see **Methods**), and then assessed the proportion of *h*^2^-cREs captured by CT-FM ICSs. Overall, CT-FM ICSs explained ∼two thirds of *h*^2^-cREs (62.5 ± 2.8% when meta-analyzed across 63 traits), which highlights the CT-FM ability to identify CTS SNP-annotations capturing most of the *h*^2^ due to known cREs (**Fig. 5e** and **Supplementary Table 19**). Of note, nearly all the *h*^2^-cREs of immune-and blood-related traits were explained by ICS SNP-annotations (84.3 ± 6.7% when meta-analyzed across 16 traits), whereas lower proportions were explained for brain-related traits (45.5 ± 3.5% when meta-analyzed across 17 traits; see **Discussion**).

We performed 6 additional CT-FM benchmarking analyses. First, we validated our estimates of *R* by performing a diagnostic procedure comparing consistency between expected and observed Z-scores ^38,58^ (mean *r*^2^ = 0.95 across the 63 GWASs, **Supplementary Fig. 17**). Second, we observed that the number of ICSs is independent of the *R* cutoff used to define an ICS (**Supplementary Fig. 18**). Third, for the 14 traits for which CT-FM identified more than one ICS, we validated that each ICS corresponds to one independent causal signal by performing S-LDSC analyses in which we jointly analyzed SNP-annotations corresponding to each ICS conditionally to each other (**Supplementary Table 20**). Fourth, we evaluated the relevance of the source of each CTS SNP-annotations included in the CT-FM model. Among the 56 high-confidence causal cell types, 33 were from ENCODE4 and 23 from CATlas. We replicated CT-FM results by leveraging CTS SNP-annotations from each of these 3 sources independently. We identified 46, 52 and 18 high-confidence causal cell types for ENCODE4, CATlas and ABC sources, respectively, which were consistent with main CT-FM results (**Supplementary Fig. 19** and **Supplementary Table 21**). These results illustrate the benefits of including CTS SNP-annotations corresponding to as many cell types and cell states as possible. Fifth, we replicated CT-FM results for 16 immune-and blood-related traits by leveraging 176 CTS SNP-annotations from EpiMap (dominated by 82 blood and immune SNP-annotations) and observed consistency across candidate cell types (**Supplementary Table 22**), which provides additional support of our primary findings. Finally, we extended our CT-FM approach to analyze 5 well-powered GWASs of East-Asian ancestry ^59^ (**Supplementary Table 23**). Despite sample size differences of these traits between East-Asian and European GWASs (average *N* = 111K vs *N* = 795K), CT-FM results were consistent across ancestries (see below and **Supplementary Table 24**), thus confirming similar CTS genetic architectures across these 2 ancestries ^60^.

Overall, CT-FM refined the number of candidate causal cell types per trait, identified highly-confident causal cell types corresponding to known cellular mechanisms, and identified ICSs with corresponding SNP-annotations explaining a large fraction of *h*^2^ captured by known cREs.

### Novel findings from traits with independent sets of causal cell types

A key feature of CT-FM compared to existing methods is to infer independent sets of causal cell types. Here, we highlight novel findings from traits for which CT-FM identified multiple ICSs (see **Fig. 5c** and **Supplementary Table 14**).

CT-FM identified 2 ICSs for height ^61^ and BMI ^62^, 2 of the most-studied complex traits. For height, CT-FM identified osteoblasts (PIP = 0.99) and fibroblasts (PIP = 0.89); these 2 cell types were also identified in a (less powered) East-Asian height GWAS (**Supplementary Table 24**). Corresponding SNP-annotations remained significant after conditioning on additional candidate fetal chondrocytes SNP-annotations ^63^, which suggests genetic mechanisms related to bone growth and connective tissues complementing those reported for embryonic cartilage cells (see **Supplementary Table 25** and **Discussion**). For BMI, CT-FM identified radial glial cells (PIP = 0.96) and bipolar neurons (PIP = 0.50), thus confirming that BMI-associated variants are likely distributed across multiple neuronal cell types ^64^. Of note, after performing S-LDSC analyses conditioning on these 2 SNP-annotations, no CTS SNP-annotations remained significant (FDR *P* < 0.05), which suggests no additional causal role of immune and adipose cell types ^16,17^ from available CTS SNP-annotations (**Supplementary Fig. 20**).

For SCZ ^65^, CT-FM detected 3 ICSs (highest number in this study), including 2 ICSs corresponding to fetal and adult excitatory neurons (PIP = 0.99 and 0.98, respectively); these 2 cell types were also inferred as candidates when analyzing a (less powered) East-Asian SCZ GWAS (**Supplementary Table 24**). Although the role of excitatory neurons in SCZ has been highlighted using gene expression datasets ^5,6^, CT-FM results suggest that different gene regulation mechanisms in excitatory neurons affect SCZ risk at early- and late-life stages (as suggested in ref. ^66^; we note that these results could also indicate the challenge of characterizing gene regulation in brain living samples, which highlights the need for both fetal and post-mortem adult brain samples ^67^). Finally, unlike in previous CTS analyses of SCZ GWAS ^4–6,65^, CT-FM could highlight the role of CD4+ T cells (PIP = 0.57), which is consistent with SCZ pathogenic mechanisms being shared with immune disorders ^68–72^. To further confirm these results, we created SNP-annotations unique to each ICS, which remained highly enriched in SCZ *h*^2^ (10.0 ± 2.7x, 12.2 ± 1.8x, and 4.5 ± 1.1x, respectively) (**Supplementary Table 26**), thereby suggesting different cellular causal mechanisms within fetal and post-mortem excitatory neurons and CD4+ T cells. Notably, although SCZ and bipolar disorder have a high genetic correlation ^73^ (*r_g_* = 0.71 +/-0.02) and both presented an adult excitatory neuron ICS, fetal excitatory neurons and CD4+ T cell ICSs were identified only for SCZ. These results suggest distinct biological mechanisms for SCZ and bipolar disorder.

Within autoimmune diseases, CT-FM identified 2 ICSs for inflammatory bowel disease ^74^ (IBD), rheumatoid arthritis ^75^ and multiple sclerosis ^76^. For IBD, the first ICS corresponded to monocyte-derived cells (max PIP = 0.63 for macrophages) and the second ICS corresponded to various types of T cells (cPIP = 0.30 and 0.45 for CD4+ and CD8+ T cells, respectively). Interestingly, when performing additional analyses of Crohn’s disease (CD) and ulcerative colitis (UC) ^74^ (2 main subtypes of IBD), CT-FM identified one of these ICSs for each subtype: monocyte-derived cell types for CD (PIP = 0.98 for macrophages) and T cells for UC (cPIP = 0.59 and 0.10 for CD4+ and CD8+ T cells, respectively) (**Fig. 5c**). Our findings are consistent with several previous reports implicating macrophages and T cells in both CD and UC ^77,78^. Notably, greatly elevated levels of monocytes were observed in patients with CD but not UC, which suggests that monocytes may play a more important role in the etiology of CD ^79,80^; conversely, several studies reported altered levels of various T-cell subsets and their effects in IBD patients as compared with healthy controls, with differences more pronounced in UC than CD patients ^81,82^. For rheumatoid arthritis, previous works have mainly characterized heritability enrichment within CD4+ T cells ^60,83,84^, but CT-FM also detected a role of variants in regions active in B cells. Indeed, B cells play critical roles in the rheumatoid arthritis pathogenesis by secreting physiologically important proteins such as rheumatoid factors, anti-citrullinated protein antibodies and pro-inflammatory cytokines ^85^. CT-FM results for an alternative rheumatoid arthritis GWAS dataset ^60^ are discussed in **Supplementary Fig. 21**. Finally, CT-FM identified 2 ICSs for multiple sclerosis, CD4+ T cells (PIP = 0.90) and macrophages (PIP = 0.99), whereas previous approaches highlighted enrichments in various immune cell types but were unable to distinguish between them ^5,6^.

Altogether, the CT-FM ability to identify independent causal cell types provided new insights into the genetic architecture of height, SCZ and multiple auto-immune diseases. CT-FM identified 2 ICSs for IBD, each corresponding to the ICS identified by CT-FM for UC and CD, which is consistent with a model of diseases being biologically heterogeneous and different disease subtypes affected by genetic risk variants via different cell types.

### Applying CT-FM-SNP to 39 UK Biobank complex traits

We applied CT-FM-SNP for 6,975 candidate causal {non-coding SNP, trait} pairs (5,819 unique SNPs) identified by SNP-fine-mapping ^32,33^ on 39 UK Biobank traits (CT-FM identified 50 ICSs across those traits; **Supplementary Table 27**). CT-FM-SNP assigned at least one ICS to nearly half (3,091/6,975; ∼44%) of the pairs, detecting a high-confidence causal cell type for most of them (2,725/3,091;∼88%) (**Fig. 6a** and **Supplementary Table 28**). The high-confidence causal cell types identified by CT-FM-SNP were consistent with CT-FM ICSs; however, CT-FM-SNP successfully nominated trait-relevant cell types for a non-negligible fraction of candidate variants (**Fig. 6b** and **Supplementary Tables 29-30**), thereby confirming the CT-FM-SNP ability to nominate causal cell types missed by CT-FM. Specifically, CT-FM-SNP linked with high-confidence mesodermal and bone marrow stromal cells to height (36 and 25 out of 405 SNPs with high-confidence cell type, respectively), mesodermal cells to bone density (29/235), mononuclear phagocytes to white blood cell count (21/89), adipocytes to waist-hip ratio (15/72), and hepatoblasts to hemoglobin A1C (9/40).

**Figure 6.**
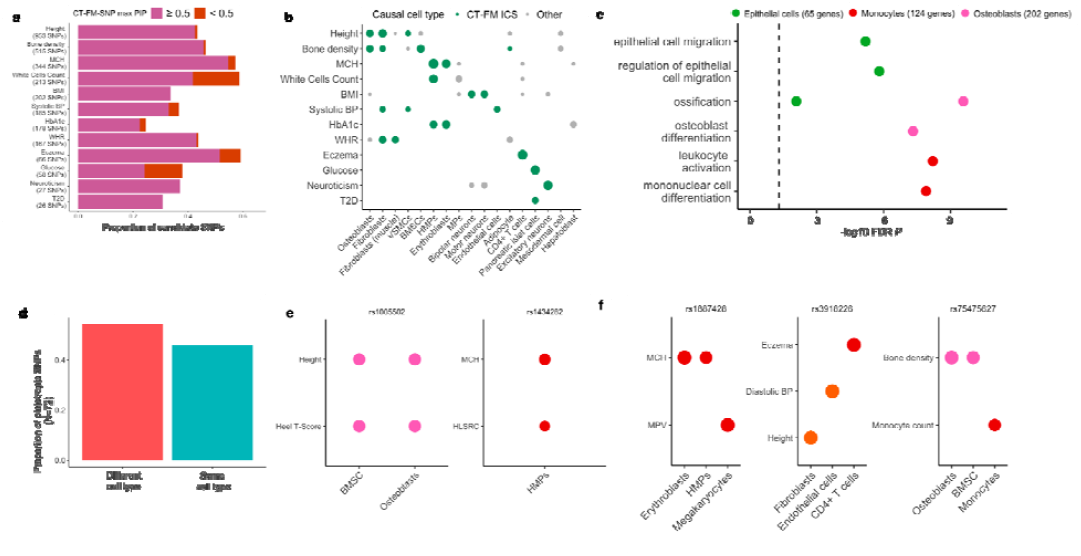
Application of CT-FM-SNP to candidate SNPs of 39 UK Biobank traits. **(a)** We report the proportion of candidate causal SNPs that were linked to at least one causal cell type by CT-FM-SNP for 12 representative traits. CT-FM-SNP results for all candidate variants of the 39 traits are reported in **Supplementary Table 28. (b)** We report the proportion of SNPs with CT-FM-SNP high-confidence causal cell type in different cell types for 9 representative traits. Cell types identified within CT-FM ICSs are represented by green dots and cell types not identified by CT-FM by grey dots. **(c)** We report the enrichment of biologically relevant processes (x axis) for genes linked to SNPs assigned to osteoblasts, epithelial cells and CD4+ T cells by CT-FM-SNP. For each process and cell type, we report the FDR-corrected enrichment p-value (y axis). Only biologically relevant gene ontology processes with FDR P < 0.01 are shown. Full gene ontology enrichment results are available in **Supplementary Table 32. (d)** We report CT-FM-SNP results for 107 pleiotropic SNPs identified across 18 genetically uncorrelated UK Biobank traits. The proportion of pleiotropic SNPs assigned to different candidate cell types is represented with a red bar, and the proportion of SNPs sharing at least 1 candidate causal cell type across traits is represented with a blue bar. CT-FM-SNP results for the 78 pleiotropic SNPs are reported in **Supplementary Table 36. (e,f)** We report 2 examples of pleiotropic SNPs assigned to the same cell type **(e)**, and 3 examples of pleiotropic SNPs assigned to different cell types **(f)**MCH: mean corpuscular hemoglobin; BMI: body mass index; BP: blood pressure; VSMCs: vascular smooth muscle cells; BMSC: bone marrow stromal cells; HLSRC: high light scatter reticulocyte count; HbA1c: hemoglobin A1c; MPV: mean platelet volume. MP: mononuclear phagocytes.

We further predicted 2,694 causal {non-coding SNP, cell type, gene, trait} quadruplets using cS2G ^39^ (**Supplementary Table 31**). To validate these predictions, we performed gene ontology enrichment analyses of genes assigned to the same cell types across the 39 traits ^86^ (**Fig. 6c** and **Supplementary Table 32**). We identified expected enrichments, such as enrichment of bone- and osteoblast-related processes for genes linked to SNPs targeting the osteoblast cell type (minimum FDR *P* = 2.6 x 10^-10^), enrichment of immune- and monocyte related processes for genes linked to SNPs targeting mononuclear phagocytes (minimum FDR *P* = 6.2 x 10^-9^), and enrichment of epithelial-related processes for genes linked to SNPs targeting epithelial cells (minimum FDR *P* = 6.5 x 10^-6^). These results underline the capacity of CT-FM-SNP and cS2G to perform predictions consistent with known biological mechanisms.

We report quadruplets consistent with known biology or highlighting new biology in **Table 1**. Specifically, for the type 2 diabetes candidate causal variant rs571342427, we inferred pancreatic islet cells and pancreatic beta cells as 2 high-confidence cell types and the insulin gene (*INS*) *^87,88^* as a target gene. For the bone density candidate causal variant rs10130587, we inferred osteoblasts as a high-confidence cell type and the bone morphogenetic protein 4 gene (*BMP4*; a gene previously implicated in body stature ^89,90^, involved in bone formation and stimulation of osteoblast differentiation ^91–93^) as a target gene. Finally, although cS2G failed to link the BMI candidate causal variant rs1421085 to *IRX3* and *IRX5* genes ^39^, CT-FM-SNP identified brain spinal cord motor neuron as a high-confidence causal cell type, thus confirming the role of the *FTO* locus in brain gene regulation ^18^. Additional details of quadruplets highlighted in **Table 1** are provided in the **Supplementary Note**.

Overall, CT-FM-SNP identified additional candidate causal cell types affecting disease risk. We further predicted 2,694 {non-coding SNP, cell type, gene, trait} quadruplets, representing one of the largest catalogs of this type to date.

### Impact of disease variants on multiple cell types

Although we identified complex traits with multiple ICSs, whether individual variants can contribute to disease risk by acting on different cell types independently is unknown. A related unanswered question is whether pleiotropic variants ^16^ influence multiple traits by acting on the same cell type or on different cell types. Here, we leveraged CT-FM and CT-FM-SNP results to tackle these questions.

To test whether traits with multiple ICSs tend to have causal variants acting jointly on multiple independent cell types, we first investigated whether the *h*^2^ explained by SNPs overlapping SNP-annotations of different ICSs was higher than expected. Specifically, for each of the 14 traits with multiple CT-FM ICSs, we ran S-LDSC with a model containing one SNP-annotation for each of the identified ICSs and one interaction SNP-annotation that exclusively contained SNPs overlapping multiple ICSs (see **Methods**). The 14 interaction SNP-annotations captured on average 1.2% of common SNPs, were expected to have a *h*^2^ enrichment of 17.3 ± 1.7x under a model with no interaction, and had an observed *h*^2^ enrichment of 16.0 ± 1.5x with the interaction (*P* = 0.22 for the conditional effect of the interaction annotation; **Supplementary Table 33**). These results suggest that, even if SNPs falling within cREs of 2 ICSs have a higher probability of being causal, these SNPs will likely affect disease risk through one of these ICSs than both ICSs.

Next, we leveraged CT-FM-SNP results to tackle this question. We focused on 6 UK Biobank traits for which CT-FM identified 2 ICSs and their corresponding candidate SNPs for which CT-FM-SNP ICSs were consistent with those of CT-FM (599 SNPs across the 6 traits; **Supplementary Fig. 22**). First, we validated that CT-FM-SNP does not overestimate the number of SNPs assigned to at least 2 CT-FM ICSs by comparing its results with *h*^2^ estimates from S-LDSC. The fraction of SNPs assigned by CT-FM-SNP to 2 CT-FM ICSs (42 ± 6% across 6 traits) was consistent with the fraction of h^2^ explained by SNPs in the intersection of CT-FM ICSs divided by the h^2^ explained by SNPs in the union of CT-FM ICSs (38 ± 8% across 6 traits) (**Supplementary Table 34**). Second, the number of SNPs assigned to both CT-FM ICSs (274) was smaller than what was expected by chance under a model in which SNPs in both CT-FM ICSs affect disease risk via a single cell type (380, more information in **Supplementary Table 35** caption). Therefore, among SNPs for which CT-FM-SNP identified 2 ICSs, it is extremely likely that most of them affect disease risk through a single ICS, which is consistent with S-LDSC and CT-FM findings above.

Finally, we investigated whether a pleiotropic SNP tends to target the same or different cell types across independent traits. We analyzed 72 candidate causal SNPs for which CT-FM-SNP identified a high confidence candidate causal cell type in at least 2 genetically uncorrelated UK Biobank traits (**Supplementary Table 36**). Approximately half of these pleiotropic SNPs (33/72; 46%) shared at least 1 candidate cell type across different traits (**Fig. 6d**). Shared {non-coding SNP, cell type} pairs across traits included SNPs acting on hematopoietic multipotent progenitors (HMPs) in blood/immune traits and SNPs acting on osteoblasts and bone marrow stromal cells in height and bone density traits (**Fig. 6e**). Strikingly, more than half of the pleiotropic SNPs (39/72; 54%) were assigned to different cell types by CT-FM-SNP (**Fig. 6f**). For example, the pleiotropic SNP rs1887428 was assigned to HMP and erythroblast cell types for mean corpuscular hemoglobin but to megakaryocytes for mean platelet volume; this result is consistent with the known role of its candidate target gene *JAK2* on regulating both erythroid progenitors ^94^ and megakaryocyte morphology ^95^. The pleiotropic SNP rs3918226 was assigned to fibroblasts, endothelial cells and CD4+ T cells for height, diastolic blood pressure and eczema, respectively; this result is consistent with the known role of its candidate target gene *NOS3* (ref. ^39^), which is produced in endothelial cells and plays a key role in regulating blood pressure ^96,97^ but was also shown to regulate CD4+ T cell production ^98,99^ and collagen synthesis in fibroblasts ^100^. Finally, the pleiotropic SNP rs75475627 was assigned to osteoblasts and bone marrow stromal cells for bone density trait and to macrophages for monocyte count trait; this result is consistent with the known role of its candidate target gene *SPTBN1*, which is involved in osteoblast differentiation and function ^101,102^ but was also shown to regulate monocyte differentiation into macrophages ^103^.

Overall, CT-FM and CT-FM-SNP results suggest that most individual SNPs affect a phenotype via a single set of cell types and that pleiotropic SNPs might target independent cell types depending on the phenotype context.

## Discussion

We developed CT-FM and CT-FM-SNP, 2 methods that leverage CTS cRE annotations to identify sets of independent causal cell types of a trait and trait-associated SNPs, respectively. By applying CT-FM to 63 GWASs, we identified 79 ICSs with corresponding SNP-annotations explaining a high fraction of trait *h*^2^, identified 14 traits with independent causal cell types, identified cell–disease relationships consistent with known biology, and highlighted previously unexplored cellular mechanisms. We applied CT-FM-SNP to 39 UK Biobank traits and predicted high-confidence causal cell types for 3,091 candidate causal non-coding SNPs-trait pairs. Finally, CT-FM and CT-FM-SNP results suggest that most individual SNPs affect a phenotype via a single set of cell types and that pleiotropic SNPs might target different cell types depending on the phenotype context.

As compared with existing methods, CT-FM and CT-FM-SNP propose several conceptual advances. First, they propose to prioritize causal cell types by leveraging cREs rather than gene expression QTL datasets ^14,19^, thus prioritizing cell types over tissues, and capturing more disease *h*^2^. Indeed, the *h*^2^ explained by the cREs identified by CT-FM is nearly 8 times higher than the *h*^2^ explained by fine-mapped eQTLs from all GTEx tissues ^21,46^ (39.0 ± 1.8% vs. 5.0 ± 0.3% across 63 traits). Second, our probabilistic framework allows for disentangling shared cREs across cell types, allowing to identify independent sets of causal cell types while greatly reducing the number of resulting candidate SNP-annotations (∼20x decrease compared to S-LDSC FDR significant results). For example, CT-FM decreased the initial association signal of S-LDSC for SCZ across 8 different biological groups to 3 independent ICSs across brain and immune cell types. Finally, CT-FM-SNP is (to our knowledge) the first method prioritizing the causal cell type of individual variants by accounting for both polygenic enrichment and cRE sharing.

Our findings have several implications for downstream analyses. First, they highlight the benefits of leveraging cREs to infer causal cell types of diseases and risk variants. Their advantages include a high fraction of *h*^2^ explained, cost-effective experiments targeting multiple cell types and conditions (ENCODE4 provides candidate *cis*-regulatory elements for 1,680 cell types and cell lines; single-cell ATAC-seq technologies now allow for inferring cREs specific to a cell state ^84^), and high potential for linking rare variants to cell types. A possible future area of research could involve developing cistrome-wide association studies ^104^ across multiple cell types using population-scale single-cell ATAC-seq data, and leverage a fine-mapping procedure (similar to refs. ^19,105^) to functionally link GWAS variants to an ATAC peak and its target cell types. Second, our application of CT-FM on 63 GWASs revealed previously unexplored cellular mechanisms and provides direction for future research. For example, although the role of chondrocytes in height has been extensively investigated ^63^, CT-FM highlighted an additional role of osteoblasts and fibroblasts in height (**Supplementary Table 25**). Third, it highlighted differences in the *h*^2^ explained by CT-FM ICSs in immune/blood- and brain-related traits. It indicated different functional architectures between those traits, a higher informativeness of functional data in immune/blood cell types that are available for a wide range of conditions (i.e., different developmental stages or *in vitro* stimulations; 552 SNP-annotations analyzed in this study), a lower informativeness of functional data in brain cell types (often collected in pre-natal or post-mortem tissues), and/or higher confounding in GWAS results of brain-related traits ^106^. Fourth, our results are consistent with a model of diseases being biologically heterogeneous and different disease subtypes affected by genetic risk variants via different cell types. For example, CT-FM identified 2 ICSs for IBD, each corresponding to the ICSs identified for CD and UC (macrophages and T cells, respectively). Identifying multiple causal cell types for a trait could allow for constructing CTS polygenic risk scores to build distinct profiles of patients ^107^. However, these results do not indicate a disease-specific action of different cell types but rather their more pronounced role in different subtypes, which involves a complex interplay between different causal cell types. Finally, CT-FM-SNP provides insights into functions of disease candidate variants and may also be helpful in choosing the relevant cell type for *in vitro* experiments.

We note several limitations of our work, related to the CT-FM and CT-FM-SNP power in inferring causation. First, CT-FM and CT-FM-SNP specifically assume that all disease-relevant cell types have corresponding SNP-annotations, but they might not have been assayed in practice. This finding is consistent with the observation that cREs are available for a wide range of immune cell types and conditions and presented the largest proportion of *h^2^-cREs* explained (**Fig. 5e**). Generating cRE catalogs of diverse cell types and conditions should fill the gap between the total trait *h*^2^ and the *h*^2^ explained by SNP-annotations from putative causal cell types. Nevertheless, CT-FM and CT-FM-SNP can still indicate the “best proxy” for the truly causal cell type. Second, as highlighted in simulations, the power to detect disease causal cell types depends on the GWAS sample size and trait *h*^2^, whose product needs to be large. Although CT-FM has been successful in detecting at least one ICS on 63 GWASs and at least 2 ICSs for 14 traits, it still might have missed some causal cell types due to lack of statistical power. However, CT-FM ICSs explained ∼two thirds of *h*^2^-cREs, which suggests that our model captures most of the causal cell types from available CTS SNP-annotations. In addition, CT-FM-SNP could detect relevant cell types that were not highlighted by CT-FM. Third, another limitation related to sample size is that we were unable to analyze GWAS of non-European ancestries, which have significantly lower sample size than GWAS of European ancestry ^108^. Specifically, we extended our CT-FM approach to analyze 31 GWASs of East-Asian ancestry ^59^ and identified only 5 traits with well-powered GWAS (**Supplementary Table 23**). Nevertheless, CT-FM confirmed similar CTS genetic architectures across these 2 ancestries ^60^ (**Supplementary Table 24**). Fourth, CT-FM power is also reduced when causal cell types have high cRE sharing. For example, in many immune-related diseases, we identified ICS related to T cells and containing both CD4+ and CD8+ T cell SNP-annotations without being able to dissociate the cases of only one or both of them being causal. Improved cRE definition, such as dynamic regulatory elements defining precise cell types or cell states ^84^, should improve the CT-FM ability to identify precise causal cell types or cell states. Finally, our CT-FM-SNP results rely on a list of candidate SNPs identified via SNP-fine-mapping and SNP-gene links from cS2G, whose powers are still limited ^32,39^.

Despite these limitations, our results demonstrate the advantage of CT-FM and CT-FM-SNP to infer independent causal cell types from GWASs, providing novel insights into the complex cell biology underlying the genetic architecture of human diseases and complex traits.

## Methods

### CT-FM method

CT-FM is a method that identifies independent sets of CTS SNP-annotations that most likely explain the *h*^2^ observed across a large set of CTS SNP-annotations. It takes as inputs GWAS summary statistics with a matching LD reference panel, and a set of CTS SNP-annotations (**Fig. 1**). Below we describe its model and parameters estimation.

We assume the infinitesimal linear model *Y* = *Xβ* + *ε*, where is a vector of quantitative phenotypes, *X* is a matrix of standardized genotypes, *β* is the vector of per-normalized-genotype effect size, and *ε* is a mean 0 vector of residuals. We model the variance of the causal effect size *β_j_* of each variant *j* as an extension of S-LDSC baseline model . Specifically, we model^4,6^.

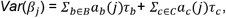

where *B* represents a set of background SNP-annotations from the baseline model (i.e., coding, enhancer, promoter etc.), *a_b_* (*j*) is the indicator variable of variant *j* for SNP-annotation *b, τ_b_*is the contribution of *b* to *Var*(*β_j_*), and *C* represents the whole set of CTS SNP-annotations. Rather than assuming that all *C* SNP-annotations contribute to *Var*(*β_j_*), we assume that at most *L* SNP-annotations contribute positively to *Var*(*β_j_*) (i.e., *τ_C_*> 0). Specifically, we model the vector *τ* of *τ*_*C*∈*C*_as 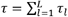 as the sum of *L* single-effects *∈_l_* = *γ_l_* · *b_l_* where *γ_l_* is a *C* ×1 binary vector indicating which CTS SNP-annotation is causal for the *l ^th^*effect, and *b_l_* is a scalar quantifying the contribution of the causal *l ^th^*effect to *Var*(*β_j_*). We caution that CT-FM specifically assumes that all disease-relevant cell types have corresponding SNP-annotations among *C*, while they might not have been assayed in practice.

In practice, we obtain marginal effects 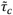 by applying S-LDSC on GWAS χ^2^ summary statistics from the following model

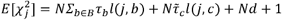

where *N* is the GWAS sample size, is a term measuring the contribution of confounding biases ^109^ and 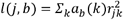 is the LD score of SNP *j* with respect to the value of SNP-annotation *a_b_*and 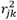 is the correlation between SNPs *j* and *k.* We can then model the vector 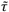 of 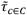 as 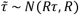, where *τ* is the sum of single effects from above, and *R* the *C* ×*C* adjusted-matrix of cRE LD scores defined as the residual correlation matrix of LD scores

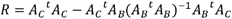

where *A_B_*and *A_C_*are the SNP × *C* and SNP × *C* SNP-annotation matrices of the LD scores of the baseline model SNP-annotations and CTS SNP-annotations, respectively. After estimating the marginal effects 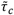 and their corresponding Z-scores using the S-LDSC framework ^29,35,36^, and computing the *R* matrix, we can estimate the number of single effects *l* ∈ *L* and corresponding *γ_l_*and *b_l_* values from the SuSiE-RSS function from the SuSiE-R package ^38^. In practice SuSiE-RSS will estimate the PIP of each SNP-annotation *c* by modeling *γ_l_*as *Multi*(1, *α_l_*), where *α_l_* is the Bayesian posterior probability of each CTS SNP-annotation to explain a single *l ^th^*effect, and by defining *PIP_c_* as

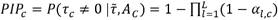

We set SuSiE parameters *L* to 10 (default) and *n* to 1,190,321 matching the number of SNPs used by S-LDSC. Of note, varying these parameters did not change the results (identical PIP values and ICSs).

We next compute -ICSs for each, where *η* represents the desired probability that the set contains causal CTS SNP-annotations (0.95 in this study). To infer an ICS for each *l*, we decreasingly sort *α_l_* and take a greedy approach to include CTS SNP-annotations until their cumulative sum exceeds *η*.

In preliminary simulation analyses, we determined that CT-FM inference procedure was well-powered when at least one out of the CTS SNP-annotations has an S-LDSC marginal 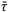 Z-score > 4 (**Supplementary Fig. 23**); we thus decided to restrict all CT-FM (and CT-FM-SNP) analyzes to summary statistics with at least one CTS SNP-annotation validating this condition (referred here as a well-powered GWAS). CT-FM then applies several steps to refine the ICS inferences. First, to remove any potential false positive ICSs, it removes ICSs in which no CTS SNP-annotations present a high S-LDSC association signal (S-LDSC Z-score > 4). Second, we introduce several modifications to the default SuSiE-RSS approach to detect potentially missed ICSs (i.e., false negatives). By default, SuSiE applies the “purity” approach, which removes ICSs whose minimum pairwise correlation *R* between its components is less than a given threshold (0.5 by default). However, this pruning method may falsely remove ICSs containing 2 causal signals exhibiting equal effect size but presenting low pairwise correlation *R*. To remedy this, we heuristically introduce a new criterion “divergence” that measures for each, the Kullback–Leibler (KL)-divergence between the distribution of *η_l_* values and the null distribution. We keep the ICSs whose purity is > 0.3 (threshold corresponding to the average absolute pairwise correlation between 2 CTS SNP-annotations in our dataset) or the KL divergence is > 3. The KL divergence of an ICS significantly correlated with the maximum S-LDSC Z-score observed in an ICS (*r*^2^ = 0.82, **Supplementary Fig. 24**), and the threshold of 3 was empirically defined because it optimizes the selection of well-powered ICSs (defined as ICSs containing at least one CTS SNP-annotation with a S-LDSC Z-score > 4). Third, in very rare cases, SuSiE-RSS can output 2 ICSs both containing a “duplicate” CTS SNP-annotation (i.e., present in both ICS1 and ICS2). To address these particular cases, we remove the ICSs containing such duplicates if the sum of their PIP values is > 0.1. In practice, only 3 detected ICSs were removed due to duplicate CTS SNP-annotations. These ICSs corresponded to systolic blood pressure (UK Biobank GWAS), forced vital capacity and the second GWAS dataset of rheumatoid arthritis (see also caption **Supplementary Fig. 21**). Finally, for ICSs for which no highly confident (PIP > 0.5) causal cell type was detected but multiple CTS SNP-annotations correspond to the same cell type, we calculate a combined PIP (cPIP) defined as:

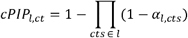

where *ct*,*l* is the cell type in a credible set *l*, and *α_l,cts_* are the *α* values of CTS SNP-annotations present in the credible set *l* and corresponding to the cell type *ct* (7 cases of cPIP > 0.5 calculated across 63 GWASs, **Supplementary Table 15**).

Finally, we note that although CT-FM generalized the SuSiE SNP-fine-mapping model, they differ in several key ways. First, CT-FM specifically assumes that all causal cell types have corresponding SNP-annotations among *C*, but they might not have been assayed in practice. Second, a *credible set* in SNP-fine-mapping is interpreted as a set of SNPs that likely contains a causal variant because in practice it is assumed unlikely that multiple SNPs in high LD are both causal. However, although mathematically identical, an ICS should be interpreted as a set of cell types that contains SNP-annotations related to one or multiple causal cell types. Indeed, distinct cell types can be causal but identified by CT-FM in the same ICS due to high cRE sharing. Thus, CT-FM results should be interpreted with some caveats in mind. For example, an ICS containing SNP-annotations corresponding to 2 distinct cell types would suggest that a) only one of these cell types is causal, b) both are causal, but their SNP-annotations are too highly correlated to be in distinct independent ICSs, or c) a third cell type that has not been assayed as causal. In addition, inferring a single cell type as causal assumes to group together several of its cell states, which does not correspond to complex disease biology.

### CT-FM-SNP method

Although CT-FM was designed to infer causal cell types underlying the overall GWAS signal, CT-FM-SNP is a method that identifies causal cell types targeted by a particular causal SNP. It takes as inputs GWAS summary statistics with a matching LD reference panel, a set of CTS SNP-annotations, and a list of candidate causal SNPs of the trait, and outputs ICSs and PIPs for those SNPs (**Fig. 1**). Briefly, for each analyzed SNP, we restrict CT-FM model to the subset of cRE annotations overlapping the SNP and perform the same inference and filtering of ICSs as described above. We additionally retain the results of SNPs that present a high-confidence causal signal (PIP > 0.5) for a well-powered CTS SNP-annotation (S-LDSC Z-score > 4) even if they were not identified in an ICS (54 such SNPs in total among 3,091 analyzed, **Supplementary Table 37**). Although the inference of multiple ICSs by CT-FM suggests that multiple cell types are causal for the disease (because it relies on polygenicity), the inference of multiple ICSs by CT-FM-SNP suggests that the candidate SNP overlap cREs from multiple causal cell types but does not infer whether the candidate SNP affects disease risk by disrupting gene regulation within a single cell type or multiple cell types.

### Selection of 924 CTS annotations for CT-FM and CT-FM-SNP

Our dataset of 924 CTS SNP-annotations (1.1% of common SNPs on average) was derived from 3 catalogs of cREs: ENCODE4, CATlas and ABC (average genome coverage = 32.5 Mb). For ENCODE4, we manually curated the dataset of 1,680 cell-type-specific cREs by primarily excluding cREs corresponding to tissues and cancer cell lines. For each resulting cRE file, we excluded marks corresponding to “*low DNase’’ and* “*Unclassified”* state. ENCODE4 data was converted from hg38 to hg19 using the *liftOver* tool from UCSC ^110^. For ABC, we retrieved a dataset of gene-enhancer predictions for 131 tissues/cell types and excluded all tissue-level annotations. CATlas data (epigenomic annotations of 222 cell types) was retrieved and converted to hg19 (liftOver) without any additional filtering. The resulting bed files were homogenized (*sortBed* function of *bedtools* ^111^) and checked for errors (*bedops --ec --merge* function of *BEDOPS ^112^*). For each dataset, we additionally created one baseline annotation corresponding to the union of all candidate regulatory regions across all cell types (see *CT-FM analyses* in the **Methods** section).

### Simulations

To assess CT-FM power under realistic scenarios, we simulated GWAS summary statistics for the 1,187,349 HapMap 3 SNPs used by S-LDSC. We considered 3 main simulation scenarios with different numbers of causal cell types: 1 (osteoblasts), 2 (osteoblasts and fibroblasts) and 3 (osteoblasts, fibroblasts and epithelial renal cells). The osteoblast and fibroblast SNP-annotations were identified on the height GWAS by CT-FM as causal (although considered independent causal cell types, these 2 SNP-annotations are non-negligibly correlated, *R* = 0.27); the epithelial renal SNP-annotation was selected according to its relatively high cRE sharing with other CTS SNP-annotations (likely due to cRE sharing with epithelial cells from other tissues; see **Supplementary Fig. 5**). We performed 500 simulations for each scenario. First, we computed expected per-SNP *h*^2^ using S-LDSC regression coefficients estimated on height GWAS with the baseline model SNP-annotations and the osteoblast SNP-annotation (when considering one causal cell type) or the osteoblast and fibroblast SNP-annotations (when considering 2 causal cell types; when considering 3 causal cell types, we modified the per-SNP *h*^2^ of SNPs in the epithelial renal SNP-annotation such that their *h*^2^ enrichment is similar to the average of *h*^2^ enrichment of SNPs in the osteoblast and fibroblast SNP-annotations). Second, we randomly selected 10,000 causal SNPs (same order of magnitude as the number of causal SNPs per GWAS in ref. ^32^), and simulated effect sizes proportional to the per-SNP *h*^2^ (total *h*^2^ was fixed at 0.5 for main simulations). Because expected per-SNP *h*^2^ can be negative, we initially set these values to 0, and rescaled positive per-SNP *h*^2^ so that expected *h*^2^ enrichment of each annotation in the model was similar to the ones observed on the height GWAS. Third, we simulated a vector of GWAS Z-scores using a multivariate normal distribution (*mvrnorm R* function), an LD matrix estimated on 337,491 unrelated British-ancestry individuals from UK Biobank release 3 (ref. ^32^), and a GWAS sample size of 350K. Fourth, we ran CT-FM using 600 CTS SNP-annotations with varying degrees of residual correlation *R* with the osteoblast and fibroblast SNP-annotations. For example, for the osteoblast cell type, we selected 40 CTS SNP-annotations presenting a weak correlation with the causal SNP-annotation (*R* between -0.11 and 0.10), 80 CTS SNP-annotations presenting a low-to-moderate correlation with the causal SNP-annotation (*R* = 0.10 to 0.32), 40 CTS SNP-annotations presenting a moderate-to-high correlation with the causal SNP-annotation (*R* = 0.32 to 0.45) and 40 CTS SNP-annotations presenting a high correlation with the causal SNP-annotation (*R* = 0.45 to 0.81). A similar selection was performed for additional 200 CTS SNP-annotations with varying degrees of correlation with the causal fibroblast CTS SNP-annotation (minimum *R* = -0.11; maximum *R* =0.78; mean *R* = 0.12) and the causal epithelial renal CTS SNP-annotation (minimum *R* = -0.14; maximum *R* = 0.56; mean *R* = 0.18) (**Supplementary Table 38**).

We performed CT-FM-SNP simulations by leveraging CT-FM simulations with one and 2 causal cell types and by sampling SNPs based on their osteoblast and fibroblast SNP-annotations and their expected per-SNP *h*^2^. We evaluated type I error within each simulation scenario by sampling 500 candidate SNPs that do not overlap the osteoblast and fibroblast causal SNP-annotations based on their expected per-SNP *h*^2^. We evaluated precision, sensitivity and F1 scores by sampling 500 candidate SNPs that overlap the osteoblast causal SNP-annotation (but not the fibroblast causal SNP-annotation) according to their expected per-SNP *h^2^.* In supplementary analyses of simulations with 2 causal cell types we also considered 500 candidate SNPs that overlap the fibroblast causal SNP-annotation (but not the osteoblast causal SNP-annotation) and 500 candidate SNPs that overlap both the osteoblast and fibroblast causal SNP-annotations.

### CT-FM analyses

We successfully ran CT-FM on GWAS summary statistics of 5 blood traits from the UK Biobank ^40^, 63 independent GWAS summary statistics, and CD and UC summary statistics ^74^ (see **Supplementary Table 5 and 14**). All GWASs were performed on individuals of European ancestry. S-LDSC Z-scores were obtained by running S-LDSC with the with the baseline v1.2 SNP-annotations; to ensure that our analyses captured CTS information, we complemented the baseline model with 6 SNP-annotations corresponding to the union of all CTS ENCODE4, CATlas, ABC, EpiMap ^25^, and DHS ^27^ cREs, as well as GTEx fine-mapped eQTLs (SuSiE PIP > 0.05 in at least one tissue) ^46,113^. We used Europeans from the 1000 Genomes Project as the reference panel ^114^.

### CT-FM-SNP analyses

We ran CT-FM on the 49 UK Biobank traits analyzed in ref. ^32^ (including the 5 blood traits analyzed in benchmarking analyses) and identified at least one ICS for 39 traits. We next leveraged functionally-informed SNP-fine-mapping results from PolyFun for these 39 traits ^32^ and identified 6,975 candidate causal {non-coding SNP, trait} pairs (5,819 unique SNPs); no CTS SNP-annotations were included in PolyFun functional priors. We applied CT-FM-SNP to all of these pairs. To link non-coding SNP to their target genes, we leveraged predictions from our cS2G strategy ^39^ and retained the gene with the highest cS2G score. GO enrichment analyses were performed using the *GOseq* R package ^86^ for cell types presenting at least 50 SNP-Cell-type pairs identified in this study. For the analyses presented in Fig. 5c and **Supplementary Table 32**, we regrouped cell types corresponding to epithelial cells (such as epithelial fibroblasts, choroid plexus epithelial cells and mammary epithelial cells). We retained all GO terms corresponding to the *Biological Process* category, containing between 10 and 1,000 genes and containing at least 5 genes from the tested gene set. All FDR significant enrichments (FDR P < 0.05) are reported in **Supplementary Table 32**. For heritability analyses presented in **Supplementary Fig. 22** and **Supplementary Tables 34-35**, we selected 6 UK Biobank traits with 2 identified CT-FM ICSs and at least 50 SNPs successfully analyzed by CT-FM-SNP. For analyses related to pleiotropic SNPs, genetically uncorrelated UK Biobank traits were defined by prioritizing UK Biobank traits with the highest SNP-heritability Z-score and by removing traits with a squared genetic correlation (*r_g_*) ^73^ > 0.05.

### OneK1K analyses

We leveraged single-cell cis-eQTLs from OneK1K ^44,45^ to functionally validate CT-FM-SNP results on lymphocyte and monocyte count candidate SNPs. We leveraged single-cell cis-eQTLs statistics computed by jaxQTL on monocytes (merging 2 OneK1K cell types – classical and non-classical monocytes) and lymphocytes (merging 4 OneK1K cell types – CD4+ T cells, CD8+ T cells, B cells, natural killer cells). For each trait, we performed cis-eQTL analyses for 3 sets of SNPs. First, we investigated lymphocyte (resp. monocyte) count candidate SNP that were assigned to lymphocyte (resp. monocyte) cell types by CT-FM-SNP. Second, we investigated lymphocyte (resp. monocyte) count candidate SNP that were not assigned to lymphocyte (resp. monocyte) cell types by CT-FM-SNP, to test whether CT-FM-SNP could successfully distinguish the candidate SNPs that do not affect trait variance by gene regulation in lymphocytes (resp. monocytes). Finally, we investigated a set of 1,393 control SNPs: we selected candidate SNPs across 16 independent UK Biobank traits not genetically correlated (*r_g_*< 0.05) to lymphocyte and monocyte count. Rather than investigating cis-eQTLs across all genes, we restricted cis-eQTL analyses to candidate genes using the PoPS method ^115^ (rank 1 to 5) on lymphocyte (resp. monocyte) count GWAS; we obtained similar trends when restricting cis-eQTL analyses to SNP-gene pairs identified by cS2G (stringent selection) and when considering all protein-coding genes (permissive selection) (**Supplementary Fig. 25**).

### S-LDSC analyses

To evaluate the *h*^2^ captured by CTS SNP-annotations identified by CT-FM, we constructed SNP-annotations representing the union of the CTS SNP-annotations within each ICS and estimated the *h*^2^ they explained using S-LDSC with baseline-LD model v2.2 and the 6 additional functional SNP-annotations used as background SNP-annotations in CT-FM analyses; we note that we followed the recommendation to use baseline model SNP-annotations for identifying critical cell types and the baseline-LD model SNP-annotations for estimating *h*^2^ enrichment ^116^. To estimate *h*^2^-cREs we merged the 6 additional functional SNP-annotations to a single annotation. Meta-analyses were performed using random-effects with the R package *rmeta*.

To investigate whether traits with independent causal cell types tend to have causal variants acting jointly on multiple cell types, we ran S-LDSC with the baseline model SNP-annotations, the 6 additional functional SNP-annotations, one SNP-annotation corresponding to each ICS and one interaction SNP-annotation corresponding to the intersection of the multiple ICSs. We meta-analyzed standardized effects □* of the interaction SNP-annotation, defined as □* = □ x *M* x sd / *h*^2^, where *M* is the number of common SNPs in the reference panel, sd is the standard deviation of the SNP-annotation, and *h*^2^ is the estimate of SNP-heritability from S-LDSC). We compared the *h*^2^ enrichment of the interaction SNP-annotation under a model with no interaction (i.e., obtained from the L estimated by S-LDSC with the baseline-LD model, the 6 additional functional SNP-annotations, and one SNP-annotation corresponding to each ICS) and with interaction (i.e., estimated directly from S-LDSC by adding the interaction SNP-annotation to the previous model).

## Data availability

S-LDSC reference files and GWAS summary statistics used in this study are available at https://zenodo.org/records/10515792. S-LDSC CTS SNP-annotations used in this study are available at https://zenodo.org/records/11194201.

## Code availability

CT-FM/CT-FM-SNP softwares and the code to replicate our analyses are available at https://github.com/ArtemKimUSC/CTFM.

## Acknowledgments

We thank G. Lettre, P. F. Sullivan, P. Dieudé, M. Vujkovic and members of the Gazal and Mancuso labs for helpful discussions. We thank the ENCODE consortium and the International Multiple Sclerosis Genetics Consortium (IMSGC) for sharing multiple sclerosis GWAS summary statistics. This research was funded by the National Institutes of Health (grant R35 GM147789).

## Competing interests

S.G reports consulting fees from Eleven Therapeutics unrelated to the present work. The other authors declare no competing interests.

## Notes

### Competing Interest Statement

Steven Gazal reports consulting fees from Eleven Therapeutics unrelated to the present work. The other authors declare no competing interests.

### Funding Statement

This research has been funded by the National Institutes of Health grant R35 GM147789.

### Author Declarations

Source data of annotations used in this study is publicly available at: http://screen-beta.wenglab.org/ https://www.engreitzlab.org/resources http://catlas.org/humanenhancer/#!/cellType The GWAS summary statistics used in this study are publicly available. The description of the GWAS files and the appropriate URL links are provided in Supplementary table 1 of this manuscript.

### Summary of Updates

Authors updated; Figures 2-6 revised; Simulation and Results sections redesigned; Reframed and re-assessed the terminology and model assumptions (Methods); Supplemental files updated

