## Supplementary Fig for "Identifying independent causal cell types for human diseases and risk variants"

### **Supplementary Note**

We discuss the {non-coding SNP, cell type, gene, trait} quadruplet predictions by CT-FM-SNP and cS2G ^1^ reported in **Table 1**.

For the asthma candidate causal variant rs479844, CT-FM-SNP inferred CD4+ T cells as a high confidence causal cell type, and cS2G inferred *OVOL1* as a target gene. While asthma risk variants linked to *OVOL1* were previously identified by GWASs ^2,3^, the role of this gene was mostly described for eczema: *OVOL1* encodes a putative zinc finger domain containing a transcription factor regulating the expression of *FLG* in atopic dermatitis and keratinocytes ^4,5^. Our predictions are consistent with the observation that *OVOL1* regulates the levels and the relative abundance of CD4+ T cells ^6,7^

For the chronotype candidate causal variant rs13081924, CT-FM-SNP inferred fetal excitatory neurons as a high confidence causal cell type, and cS2G inferred *WNT7A* as a target gene. *WNT7A* is a member of the conserved Wnt signaling pathway previously implicated in the regulation of sleep cycles ^8,9^. It is also a key regulator of brain development ^10^ and specifically regulates the number and strength of excitatory synapses (consistent with CT-FM-SNP result), with a less pronounced role in inhibitory neurons ^11^.

For the lymphocyte count candidate causal variant rs35592432, CT-FM-SNP inferred CD8+ T cells as a high confidence causal cell type, and cS2G inferred *FOXP1* as a target gene. *FOXP1* is an ubiquitously expressed gene implicated in various biological processes including development and regulation of various immune cells ^12,13^. It is also implicated in T cells regulation, in particular by regulating CD8+ T cells quiescence ^14^ (consistent with CT-FM-SNP result). Additionally, a more pronounced impact of *FOXP1* deletion on CD8+ T cells rather than CD4+ T cells was reported ^15,16^.

For the neuroticism candidate causal variant rs34272688, CT-FM-SNP inferred fetal excitatory neurons as a high confidence causal cell type, and cS2G inferred *ATAD2B* as a target gene. Association between *ATAD2B* variants and neuroticism was previously reported in a GWAS ^17^, and the transient expression of *ATAD2B* was reported in developing neurons ^18^ (consistent with CT-FM-SNP result).

For the platelet count and platelet volume candidate causal variant rs117672662, CT-FM-SNP inferred megakaryocytes as a high confidence causal cell type, and cS2G inferred *ACTN1* as a target gene. *ACTN1* is mainly expressed in megakaryocytes and mature platelets^19^ and *ACTN1* mutations were previously implicated in macrothrombocytopenia - a rare condition associated with lower platelet counts and abnormally large platelets ^20^ (consistent with CT-FM-SNP result).

Finally, for the platelet volume candidate causal variant rs998908, CT-FM-SNP inferred megakaryocytes as a high confidence causal cell type, and cS2G inferred *CD9* as a target gene. *CD9* belongs to a family of tetraspanins and is involved in megakaryocyte differentiation ^21,22^ (consistent with CT-FM-SNP result).

### **Supplementary Figures**

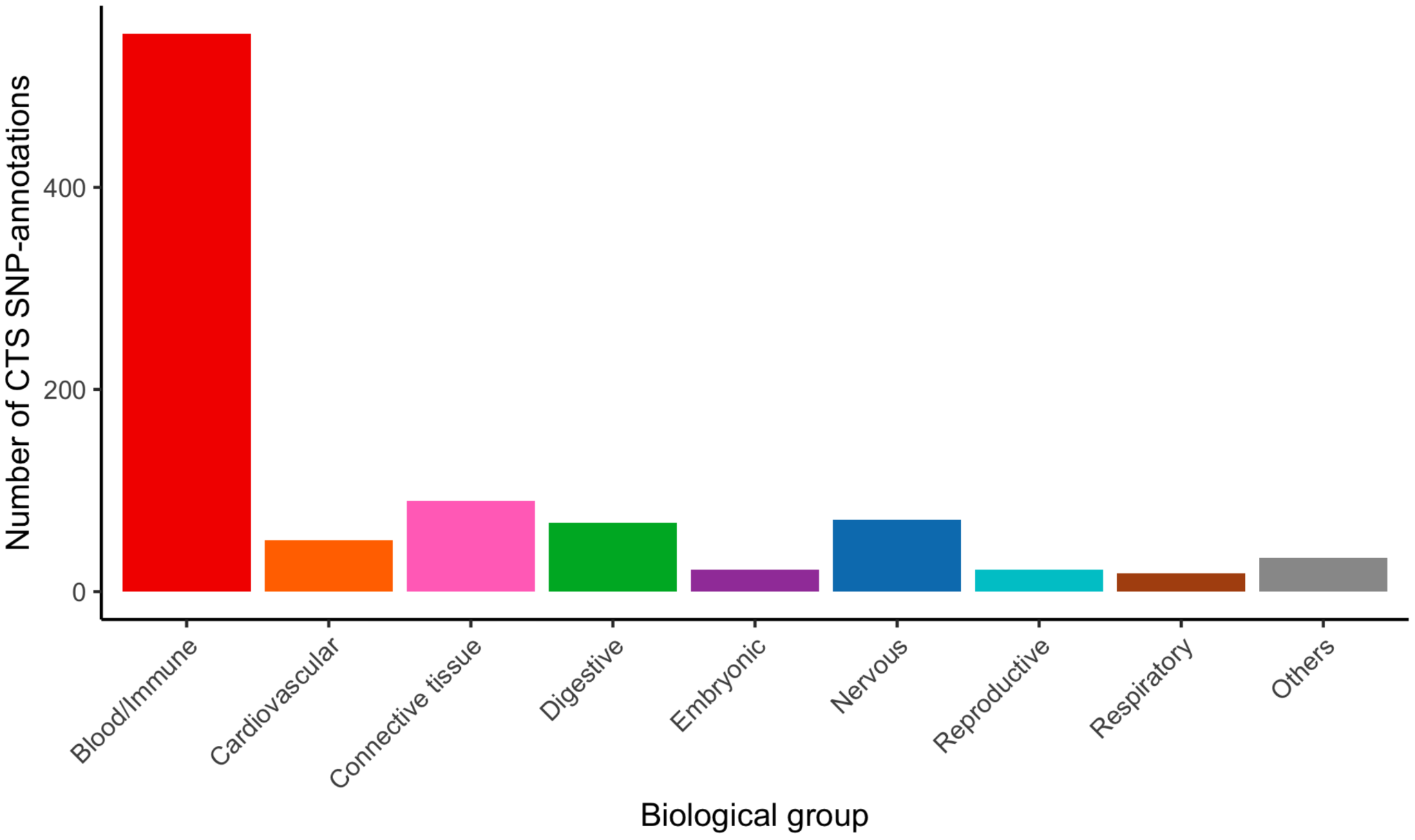

**Supplementary Figure 1. Overview of CTS SNP-annotations used in this study.** We report the number of CTS SNP-annotations within each biological group. A total of 924 CTS SNP-annotations were retrieved from ENCODE4 (650), ABC model (52) and CATlas (222) and assigned to one of the nine biological groups.

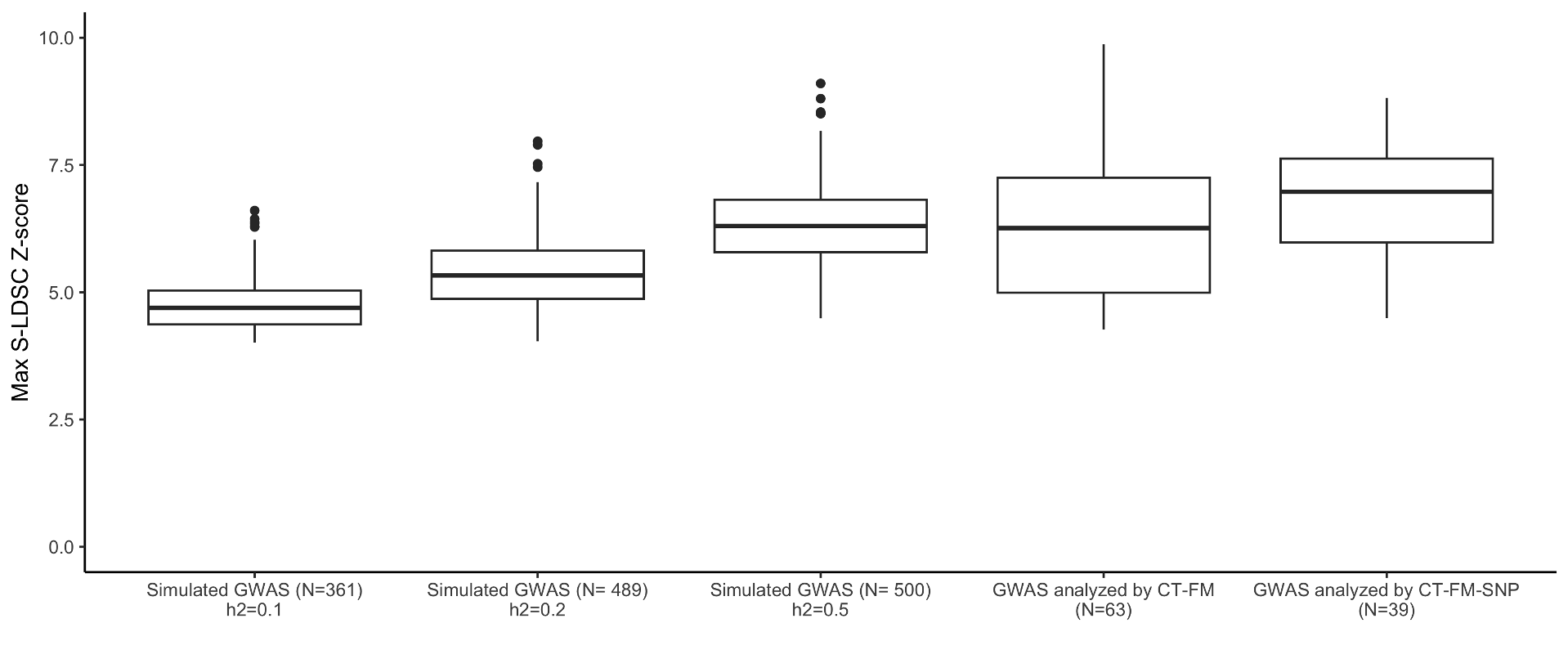

**Supplementary Figure 2. Maximum S-LDSC Z-scores obtained in simulations and real data.** We report boxplots of the maximum S-LDSC Z-score obtained across 3 simulation settings with different *h^2^* parameter (we restricted analyses to simulations with maximum S-LDSC Z-score > 4), across the 63 independent GWASs analyzed by CT-FM, and the 39 UK Biobank GWASs analyzed by CT-FM-SNP. We observed that GWASs simulated by fixing *N* = 350K and *h^2^* = 0.5 provided a median of maximum S-LDSC Z score similar to those observed in the GWAS analyzed in this study.

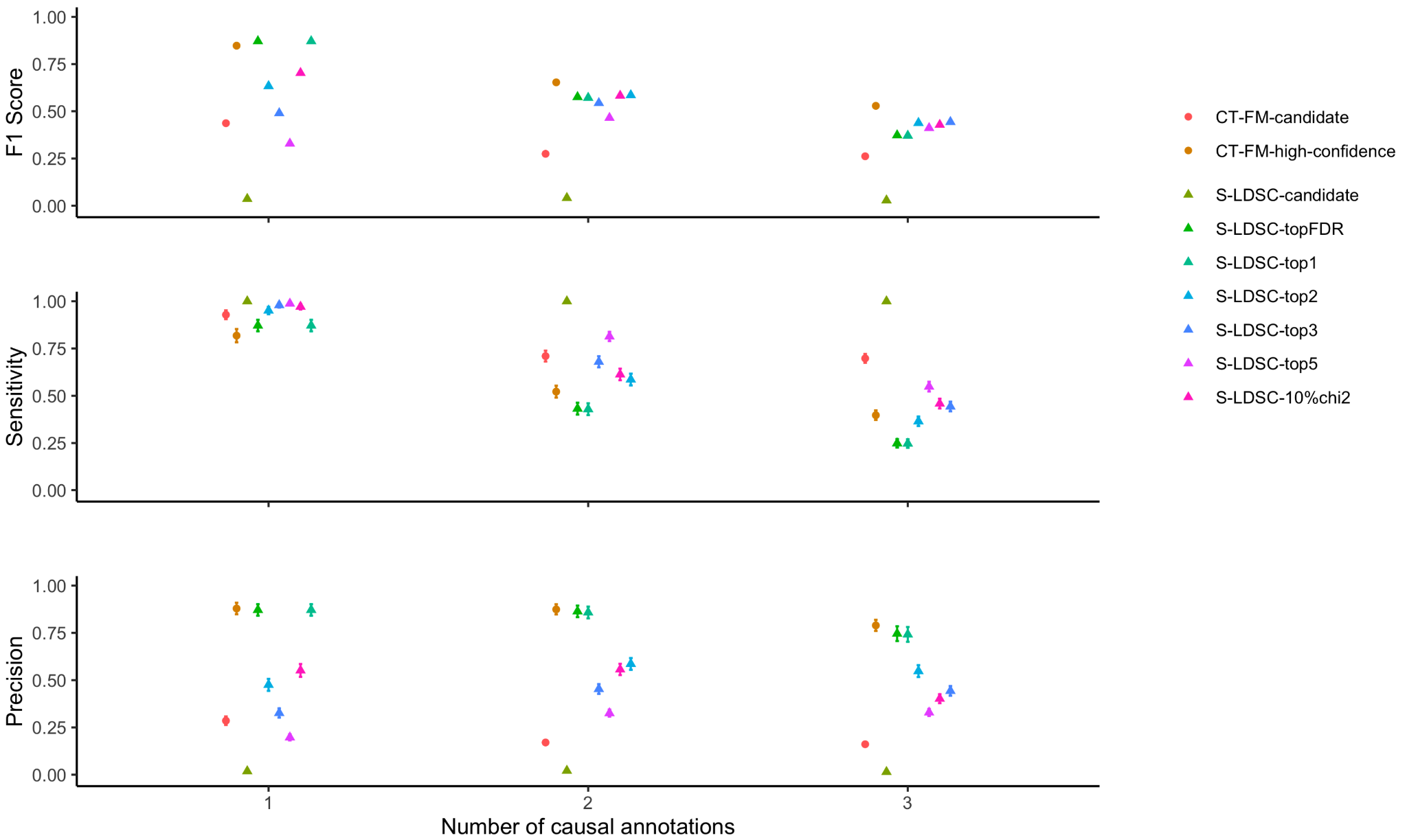

**Supplementary Figure 3. Simulations to assess precision and sensitivity of additional approaches based on S-LDSC FDR *P*.** We report the precision, sensitivity and F1 score in simulations with different numbers of causal annotations. We evaluated seven approaches based on S-LDSC FDR *P*: S-LDSC-candidate (reporting SNP-annotations with S-LDSC FDR *P* < 0.05), S-LDSC-topFDR (reporting the SNP-annotation with the most significant S-LDSC FDR *P*), S-LDSC-top1 (reporting the SNP-annotation with the smallest S-LDSC *P*), S-LDSC-top2 (reporting the 2 SNP-annotations with the smallest S-LDSC *P*), S-LDSC-top3 (reporting the 3 SNP-annotations with the smallest S-LDSC *P*), S-LDSC-top5 (reporting the 5 SNP-annotations with the smallest S-LDSC *P*) and S-LDSC-10%chi2 (reporting the SNP-annotations with chi-square that is within 10% of the lead SNP-annotation). Overall, we observed that S-LDSC-topX nearly maximized F1 score when considering X causal annotations, while CT-FM-high-confidence maximized F1 score when >1 causal annotations. Error bars represent 95% confidence intervals.

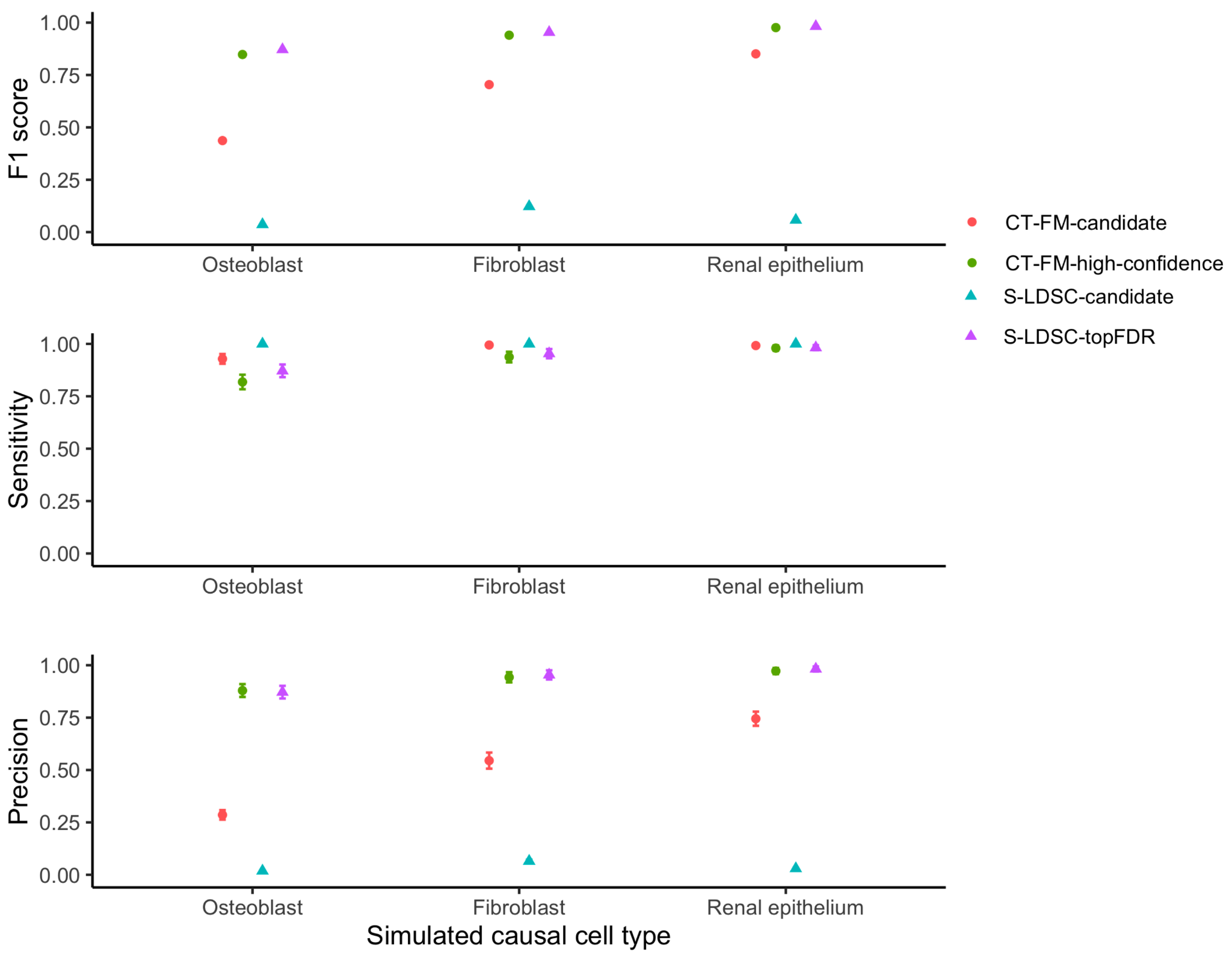

**Supplementary Figure 4. Simulations to assess precision and sensitivity of methods inferring causal cell types using different causal cell types.** We report the precision, sensitivity and F1 score in simulations with different single causal annotations. We observed nearly similar results for CT-FM-high-confidence. We observed that precision and sensitivity were decreasing when the causal SNP-annotation had high value *R* (i.e., correlation of the LD scores adjusted on the background SNP-annotations) with other analyzed SNP-annotations (see **Supplementary Figure 5**). When using osteoblast and fibroblast as the causal cell type, we performed simulations using per-SNP *h^2^* estimated on height with

Error bars represent 95% confidence intervals.

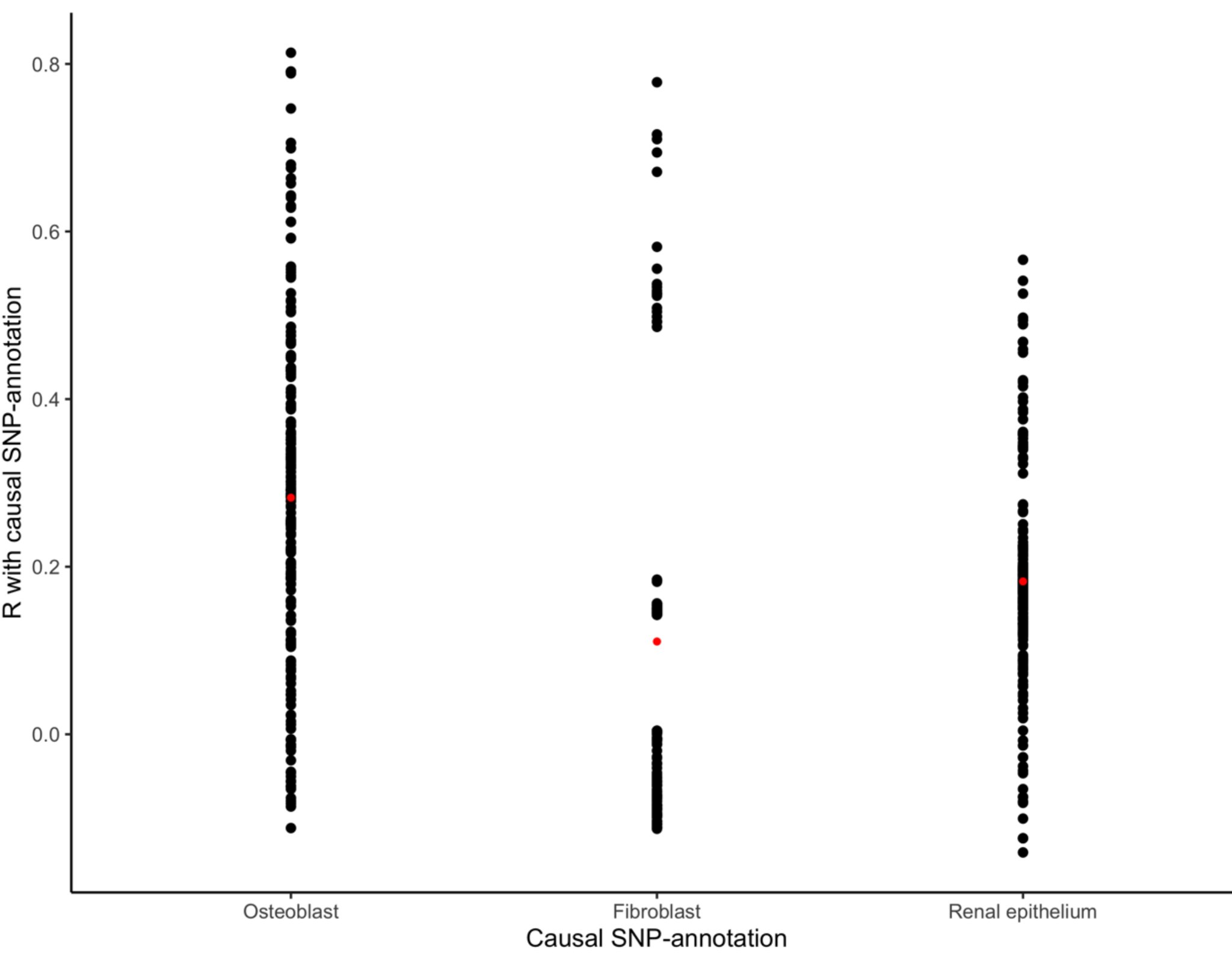

**Supplementary Figure 5. *R* values between causal SNP-annotations and other analyzed SNP annotations.** We report *R* (i.e., correlation of the LD scores adjusted on the background SNP-annotations) of the three SNP-annotations considered as causal in simulations with other analyzed SNP-annotations. We observed that the osteoblast SNP-annotation is highly correlated (i.e., *R* > 0.5) with a lot of other analyzed SNP-annotations, while the renal SNP-annotation is highly correlated for a limited number of annotations. The red dots correspond to the mean value of *R*.

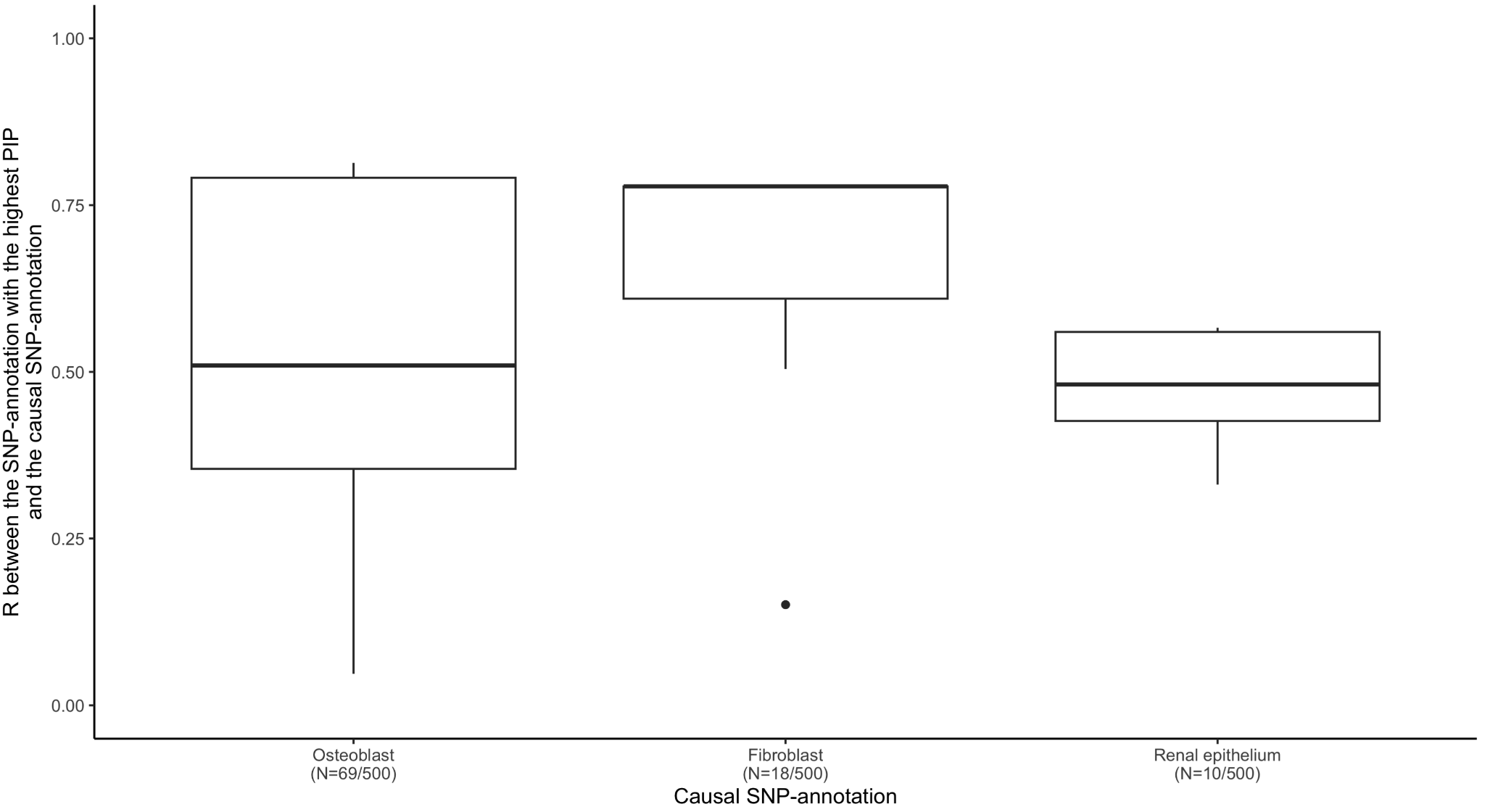

**Supplementary Figure 6. *R* values between the SNP-annotation with the highest CT-FM PIP and the causal SNP-annotation in unsuccessful CT-FM iterations.** For simulations with 1 causal cell type, we observed 69 (resp. 18 and 10) iterations in which CT-FM did not attribute the highest PIP to the causal osteoblast (resp. fibroblast and renal) CTS-SNP annotation.

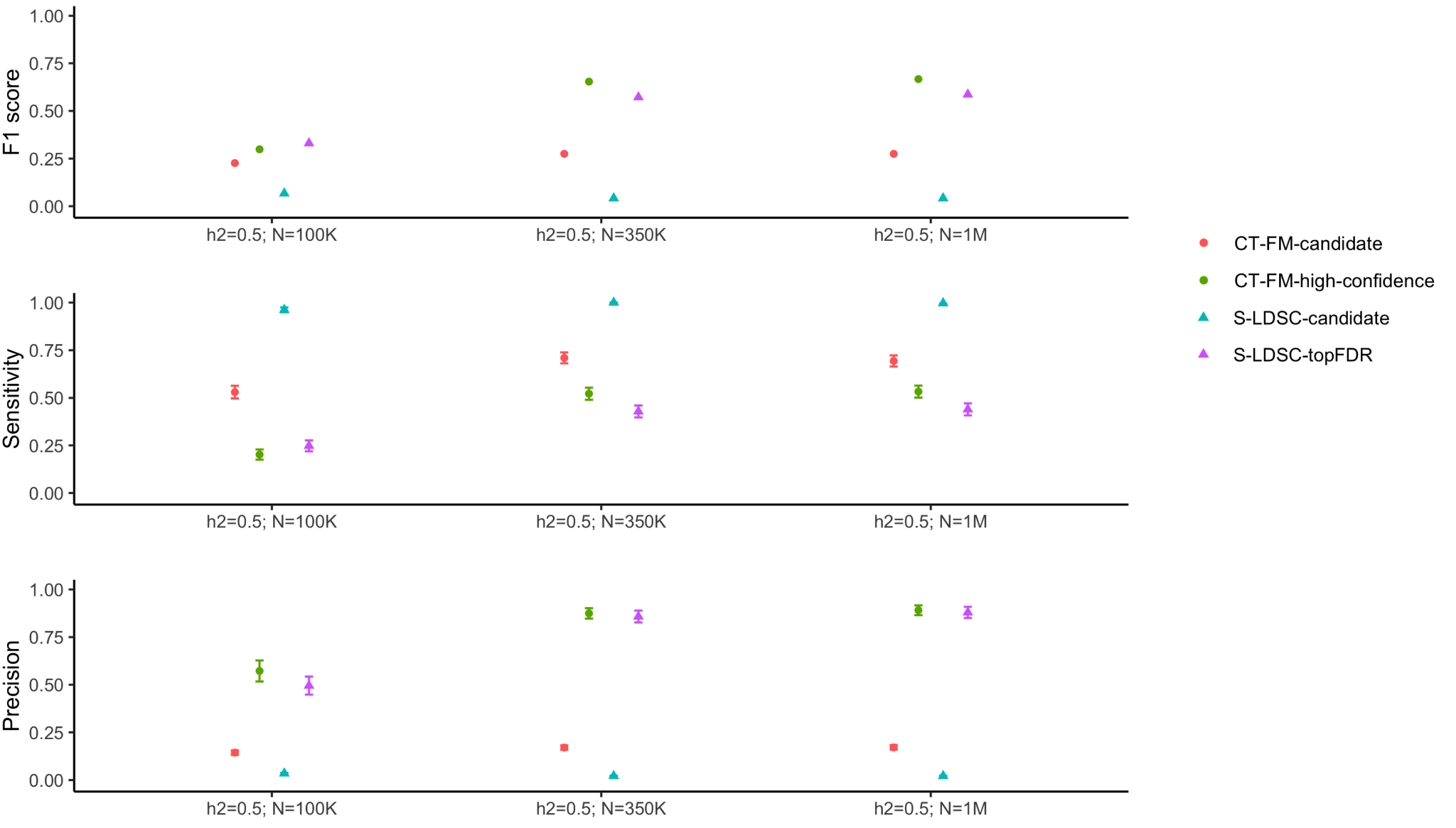

**Supplementary Figure 7. Simulations with different sample sizes (*N*) to assess precision and sensitivity of methods inferring causal cell types.** We report the precision, sensitivity and F1 score in simulations with two causal SNP-annotations where we let vary *N*. Error bars represent 95% confidence intervals.

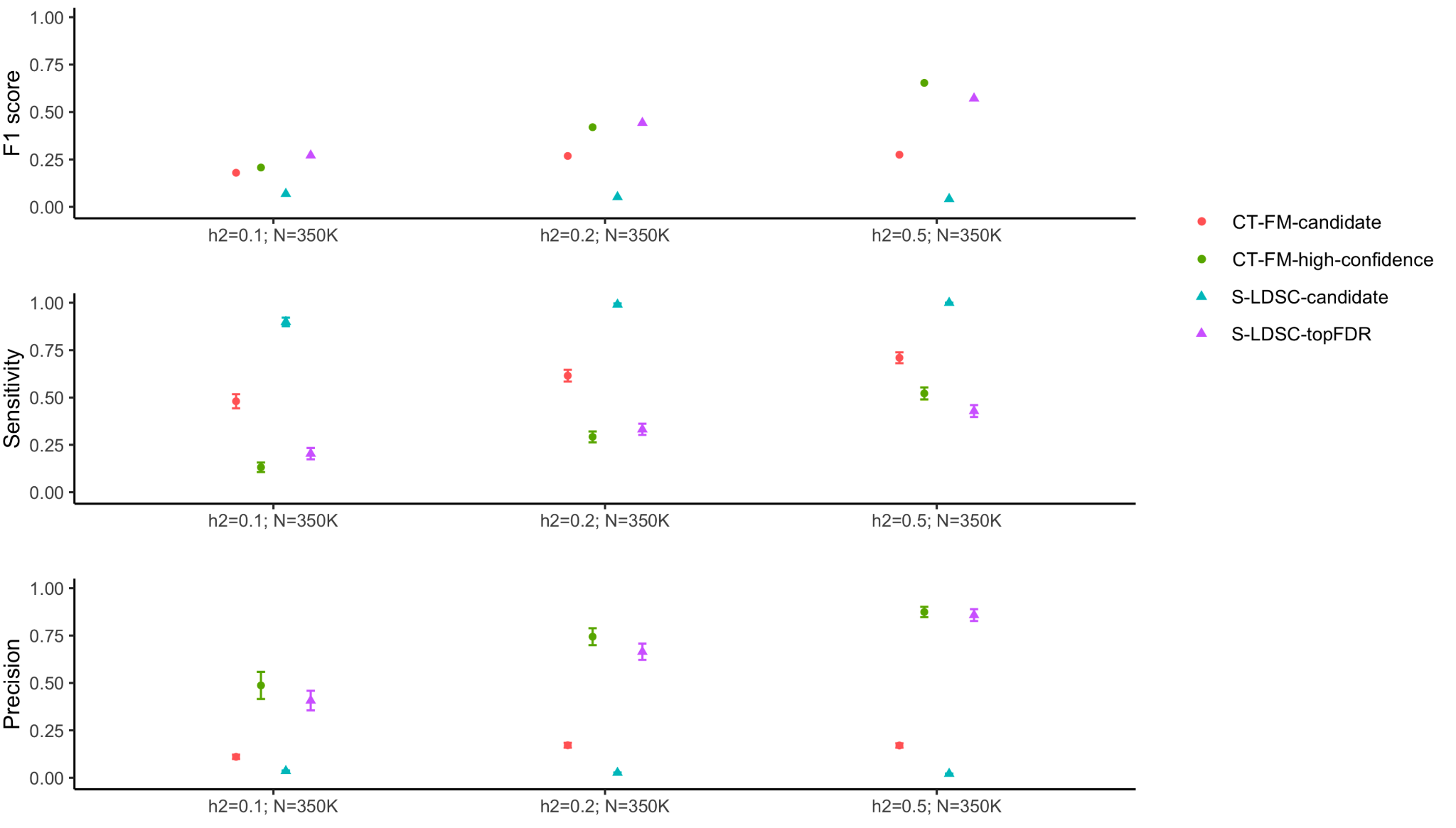

**Supplementary Figure 8. Simulations with different *h^2^* to assess precision and sensitivity of methods inferring causal cell types.** We report the precision, sensitivity and F1 score in simulations with two causal SNP-annotations where we let vary *h^2^*. Error bars represent 95% confidence intervals.

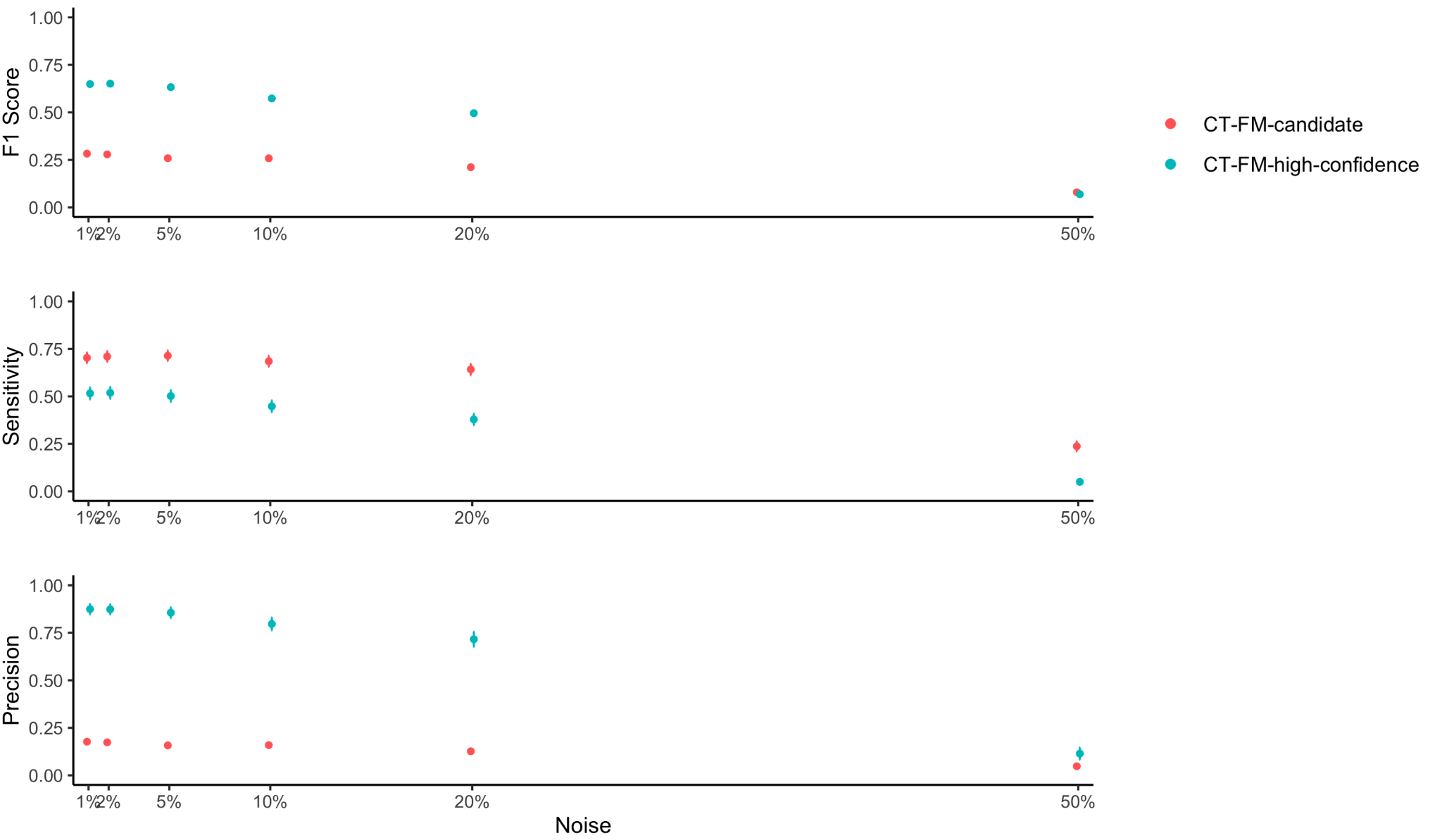

**Supplementary Figure 9. Simulations adding noise to the causal SNP-annotation.** We report the precision, sensitivity and F1 score in simulations where we added noise in the two causal SNP-annotations (i.e., changing X% of 1 into 0 in the causal SNP-annotations, and a similar number of 0 into 1, for different values of X). Error bars represent 95% confidence intervals.

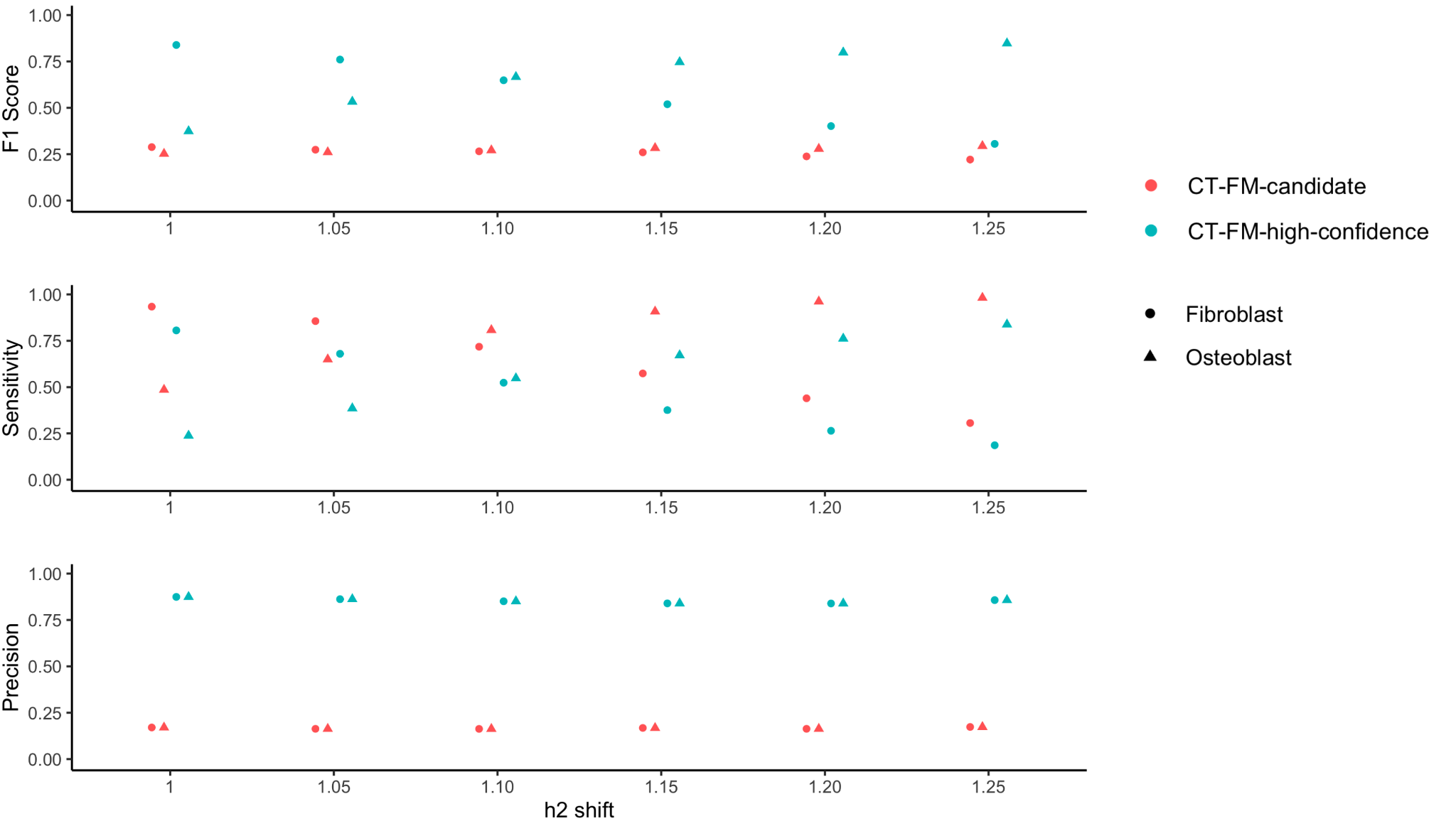

**Supplementary Figure 10. Simulations where the osteoblast and fibroblast SNP-annotations contribute inequitably to heritability.** We report the precision, sensitivity and F1 score in simulations where the osteoblast and fibroblast SNP-annotations contribute inequitably to heritability. Specifically, for different values of X (x-axis) we divided the *h^2^* explained by the osteoblast SNP-annotation by X, and multiplied the *h^2^* explained by the fibroblast SNP-annotation by X. We observed that precision remained stable, while sensitivity decreased (resp. increased) when the SNP-annotations explained less (resp. more) *h^2^*. The average F1 score across the 2 causal annotations remained similar across X. Error bars represent 95% confidence intervals.

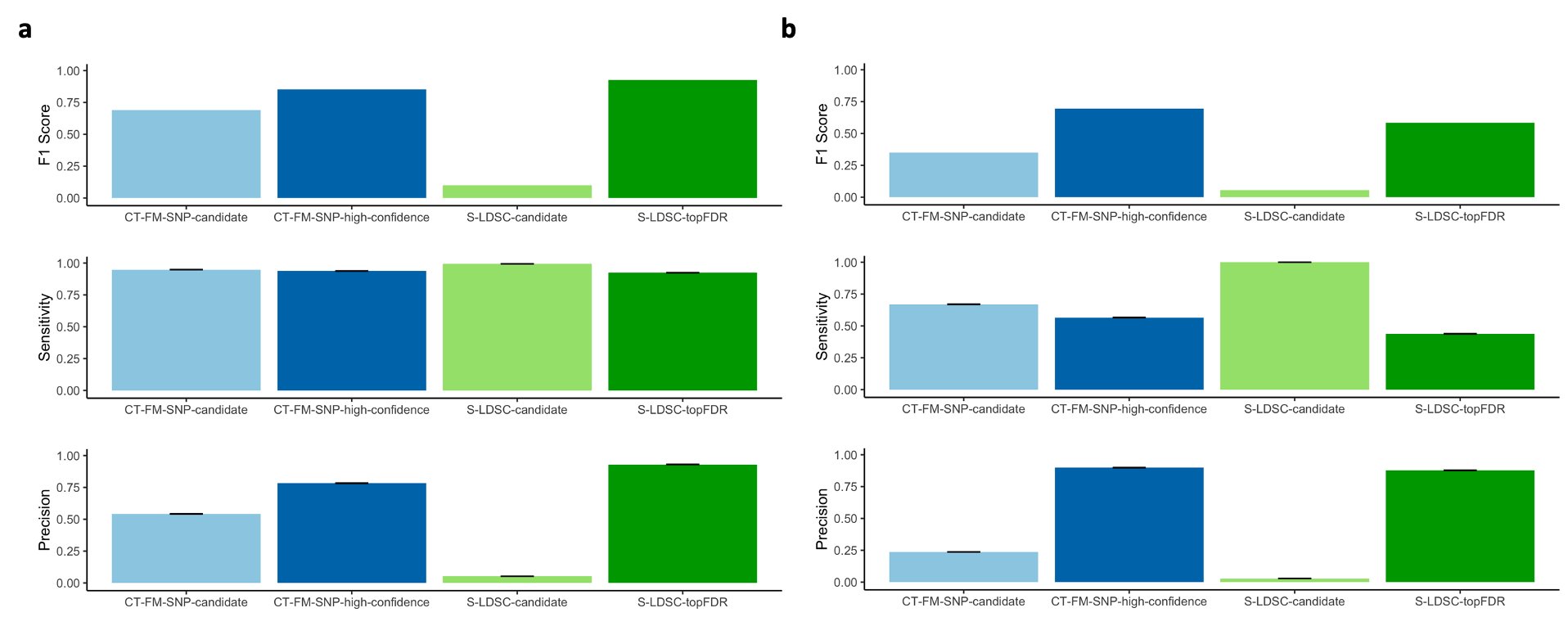

**Supplementary Figure 11. Simulations to assess precision and sensitivity of methods inferring causal cell types of candidate SNPs.** We report the precision, sensitivity and F1 score in simulations where we considered osteoblast and fibroblast as causal cell types. We report results for candidate SNPs overlapping the fibroblast SNP-annotation in (**a**), and for candidate SNPs overlapping both osteoblast and fibroblast annotations in (**b**). Error bars represent 95% confidence intervals.

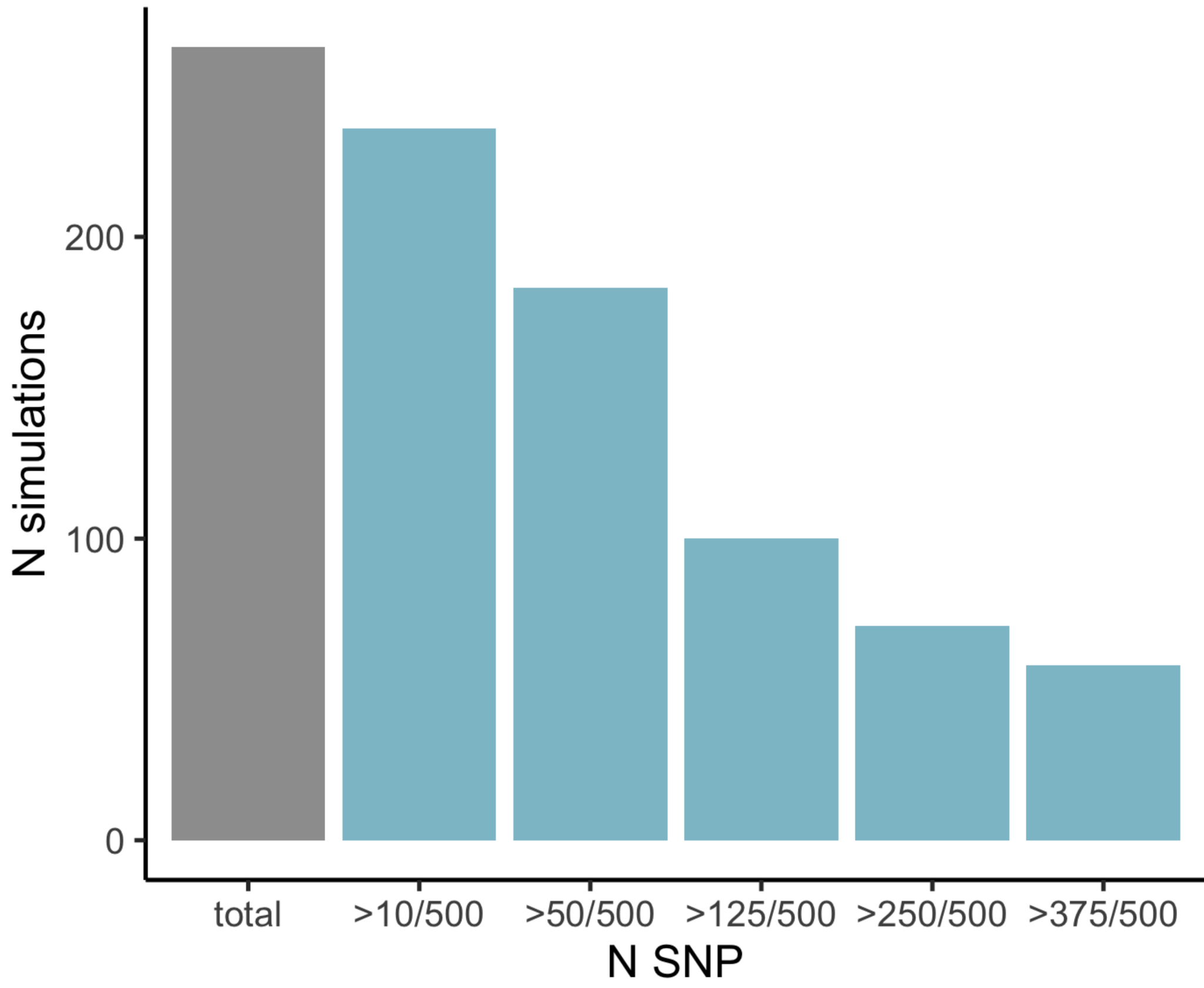

**Supplementary Figure 12. Number of SNPs assigned to osteoblasts by CT-FM-SNP in simulations where CT-FM did not identify osteoblast as a causal cell type.** In simulations with osteoblast and fibroblast as the causal cell types, we observed 263/500 simulations where CT-FM did not identify osteoblast in an ICS. For these simulations, we observed that CT-FM-SNP was able to assign >50 (resp. 125 and 250) of the 500 candidate SNPs to the osteoblast annotation with high confidence in 78% (resp. 38% and 27%) of the cases.

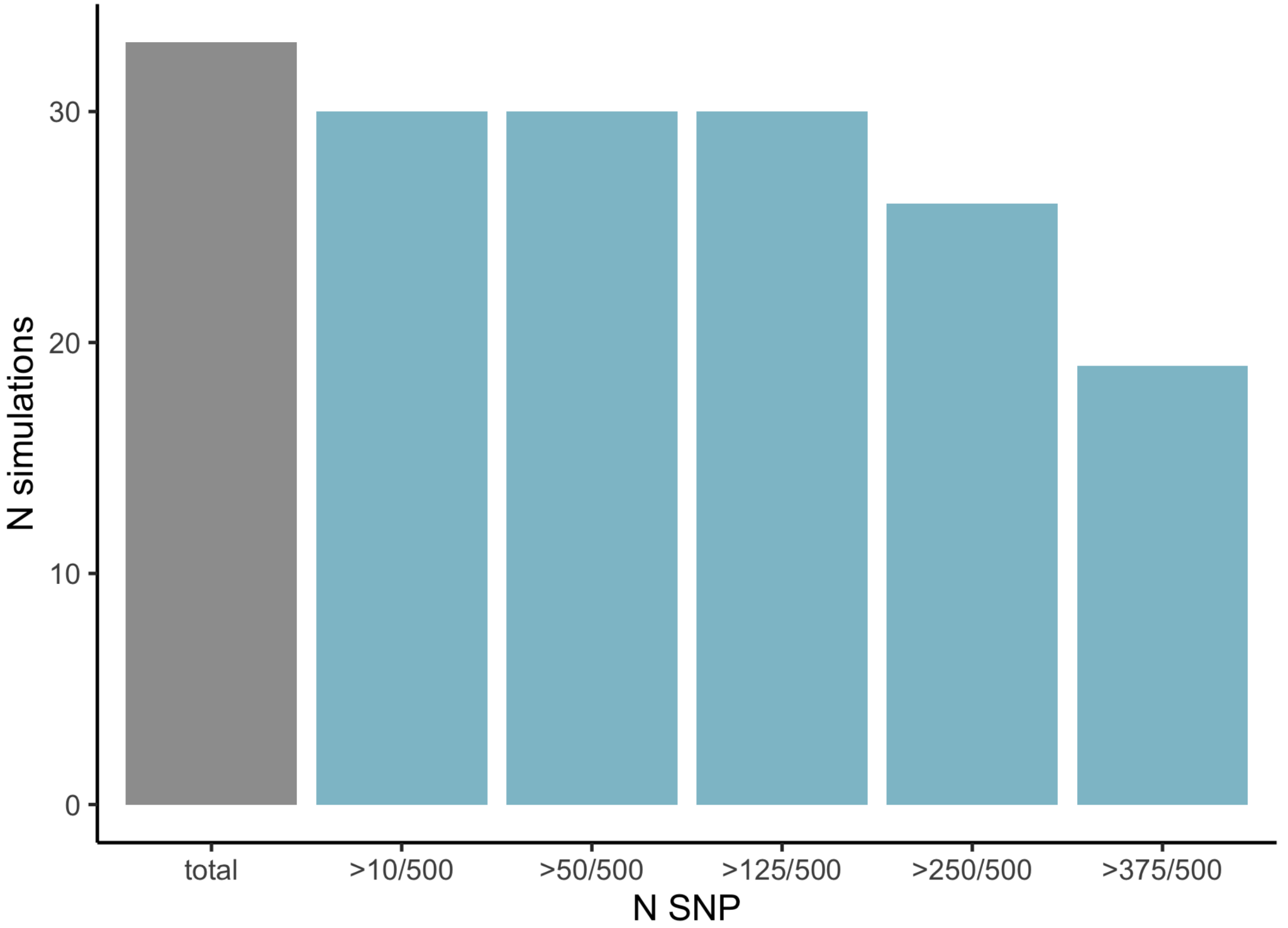

**Supplementary Figure 13. Number of SNPs assigned to fibroblasts by CT-FM-SNP in simulations where CT-FM did not identify fibroblast as a causal cell type.** In simulations with osteoblast and fibroblast as the causal cell types, we observed 33/500 simulations where CT-FM did not identify fibroblast in an ICS. For these simulations, we observed that CT-FM-SNP was able to assign >50 (resp. 125 and 250) of the 500 candidate SNPs to the fibroblast annotation with high confidence in 91% (resp. 91% and 79%) of the cases.

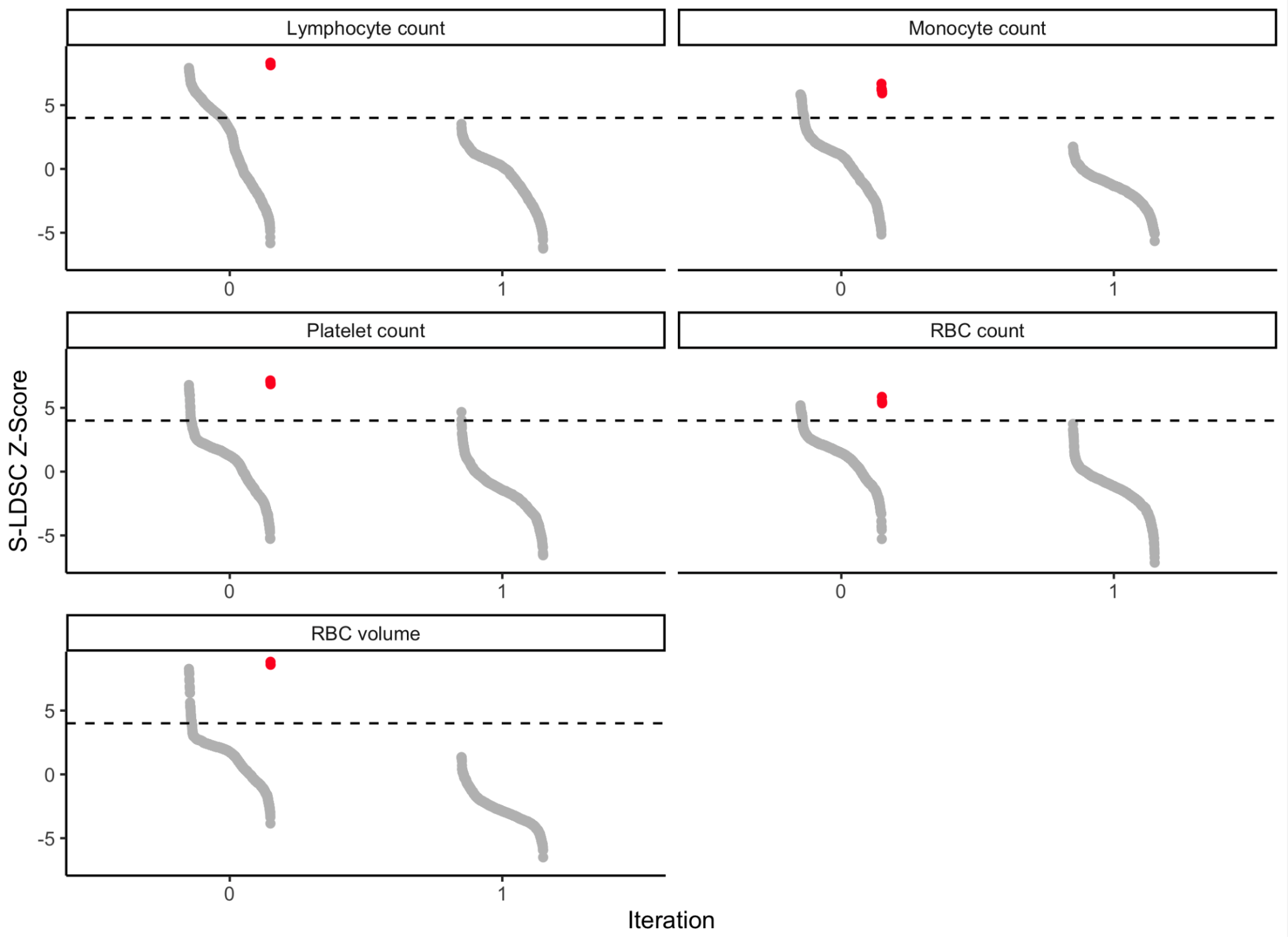

**Supplementary Figure 14. Conditional S-LDSC analysis for five blood traits.** We report the initial S-LDSC Z-scores of the 924 CTS SNP-annotations used by CT-FM under iteration 0. Candidate causal CTS SNP-annotations (i.e., assigned to a CS by CT-FM) are indicated in red. To validate that the CSs detected by CT-FM captured most of the conditionally independent causal signal, we reran S-LDSC conditioned on the SNP-annotations in CT-FM CSs (and the background annotations) on the remaining CTS SNP-annotations (i.e., not assigned to a CS by CT-FM). We report the new S-LDSC Z-scores under iteration 1. For 4 out of the analyzed 5 traits, we observed that none of the remaining CTS SNP-annotations had a $\tilde{\tau}$ Z-score > 4, confirming that no CTS conditional effect remains in the data after identifying CS with CT-FM. For platelet count, 2 of the remaining 919 CTS SNP annotations presented a $\tilde{\tau}$ Z-score > 4 after conditional S-LDSC analysis, corresponding to bone marrow myeloid progenitors (Z = 4.02, similar CTS SNP annotations were previously identified in a CT-FM CS) and fetal megakaryocytes (Z = 4.65), for which CT-FM-SNP assigned a substantial number of candidate SNPs (see main text). The dashed horizontal line represents S-LDSC $\tilde{\tau}$ Z-score = 4.

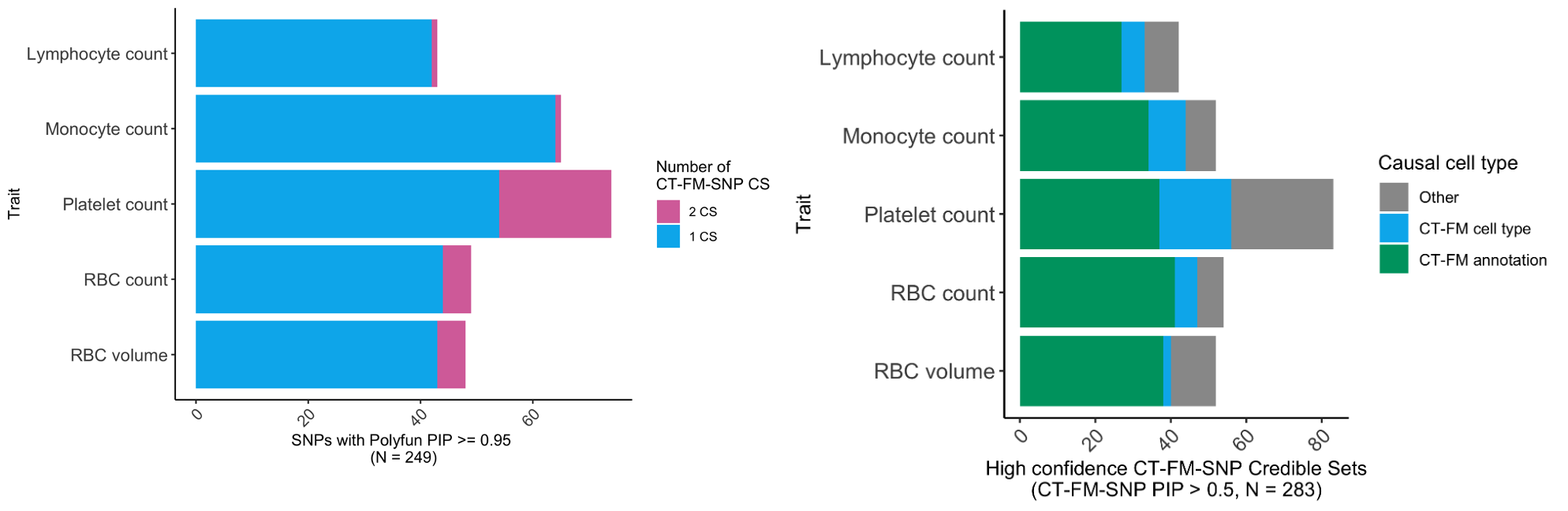

**Supplementary Figure 15. CT-FM-SNP results in five blood cell traits when restricting analyses to 249 SNPs with Polyfun SNP-PIP > 0.95. (left)** We report the number of candidate causal SNPs that were linked to at least one causal cell type by CT-FM-SNP. CT-FM-SNP results for all candidate variants are reported in **Supplementary Table 8**. **(right)** We report the number of high confidence {non-coding SNP, cell type, trait} triplets inferred by CT-FM-SNP where the cell type is consistent with CT-FM results. We highlight triplets where the causal CTS SNP-annotation was also found in CT-FM CSs (green), triplets where the causal CTS SNP-annotation was not found in CT-FM CSs, but corresponds to the same cell type (blue), and triplets where the causal CTS SNP-annotation was not found in CT-FM CSs (grey). Overall, we observed high consistently between CT-FM-SNP analyses performed on candidate SNPs with SNP-PIP > 0.95 (main **Fig. 4**) and SNP-PIP > 0.95, demonstrating that conclusions from our main analyses are robust to the imperfect selection of candidate causal SNPs. RBC: red blood cell.

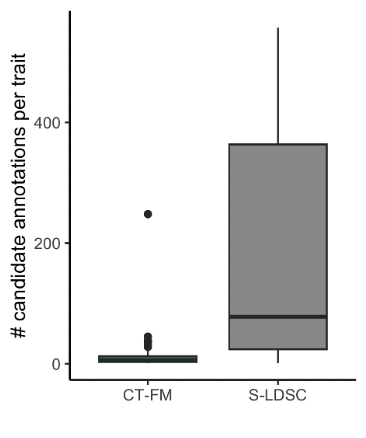

**Supplementary Figure 16. S-LDSC and CT-FM results for 63 traits** ^60^**.** We report the distribution of candidate causal CTS SNP-annotations inferred by CT-FM and S-LDSC. The median value of confident scores is displayed as a band inside each box; boxes denote values in the second and third quartiles; the length of each whisker is 1.5 times the interquartile range, defined as the width of each box. Numerical results are reported in **Supplementary Table 16**.

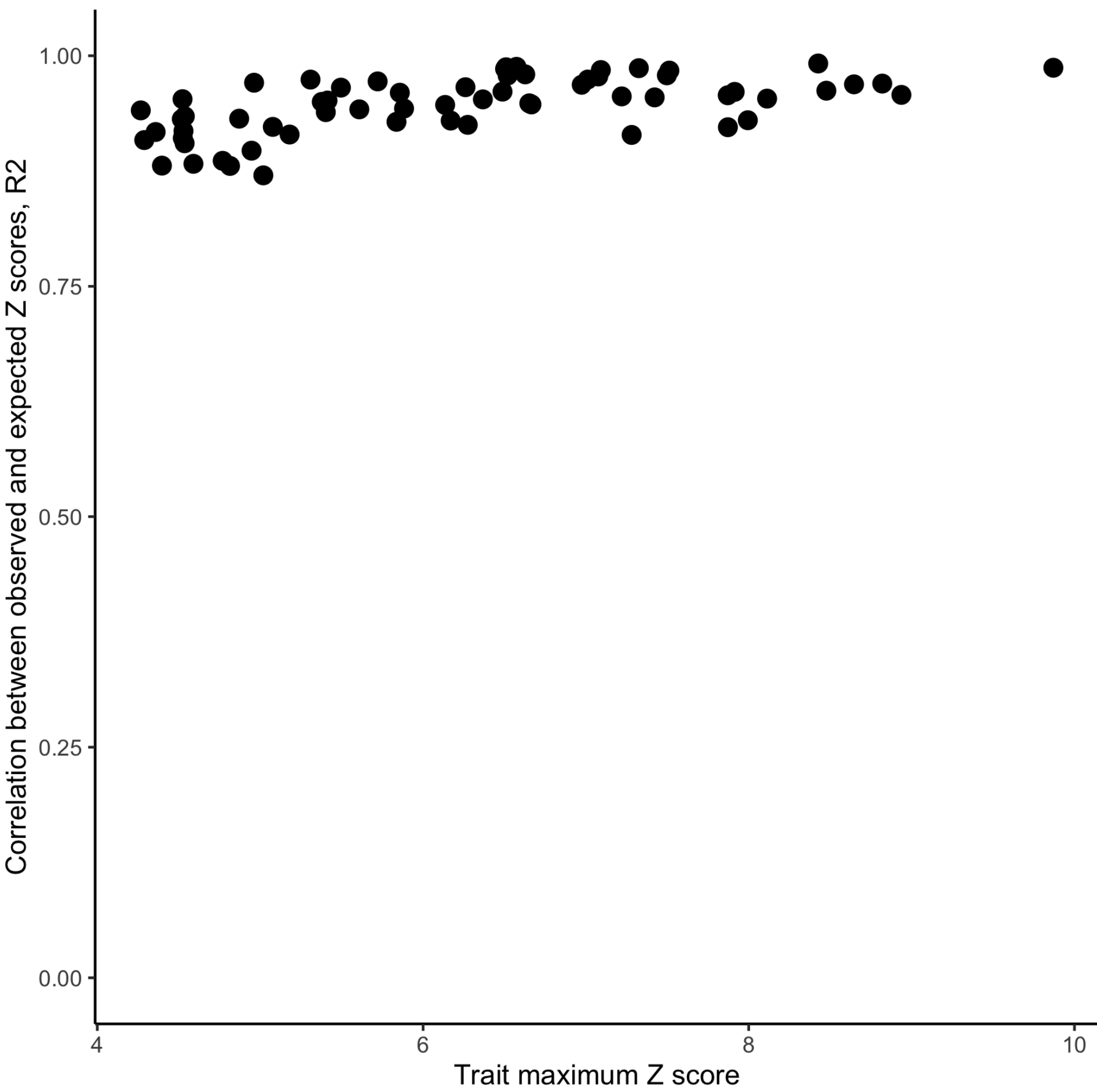

**Supplementary Figure 17. Concordance between expected and observed Z-scores across 63 GWASs.** We report the correlation coefficients *r*^2^ between observed and expected Z scores calculated by SuSiE-RSS kriging approach ^24,25^. We observed a high concordance between expected and observed Z-scores across traits (mean *r*^2^ = 0.95 across the 63 GWASs) indicating reliability of CT-FM fine-mapping results.

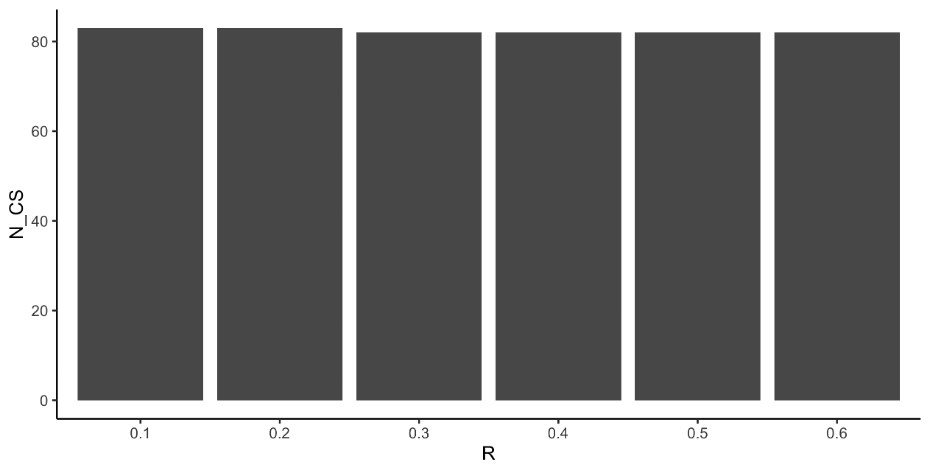

**Supplementary Figure 18. The number of ICSs per trait is independent of the *R* cutoff used to define an ICS**. We report the number of ICS identified for 63 traits (y axis) when using different values for R cutoff (*min_abs_corr* parameter in SuSiE, x axis).

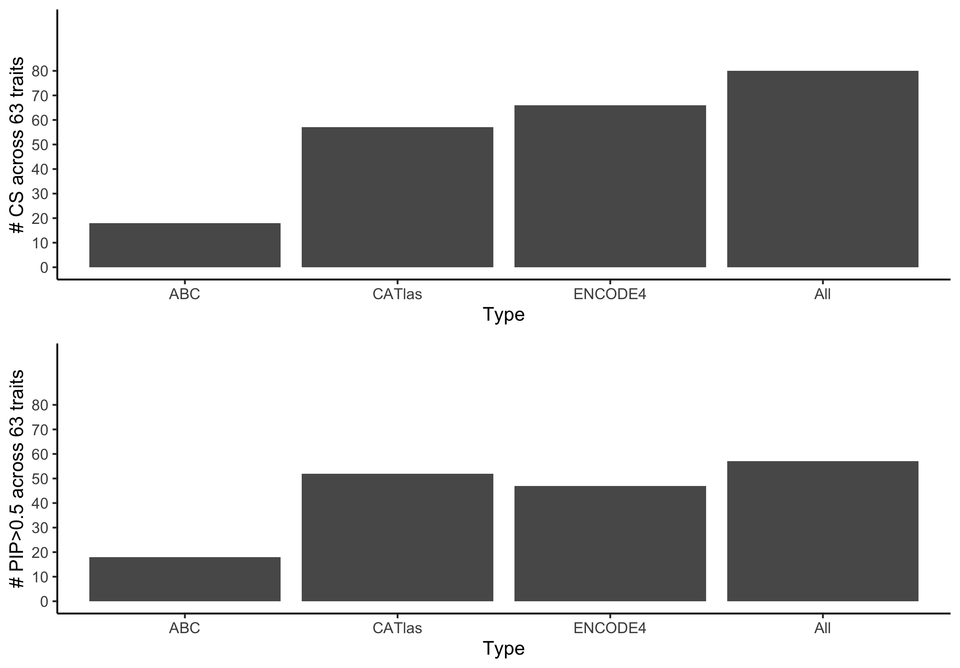

**Supplementary Figure 19. CT-FM results when using CTS SNP-annotations from a single source.** We report CT-FM results (number of credible sets and number of high confidence PIP>0.5 causal CTS SNP-annotations) obtained when using exclusively ABC (52 annotations), CATlas (222 annotations) and ENCODE4 (652 annotations) datasets. We identified 66, 57 and 18 CSs and 46, 52 and 18 high-confidence causal cell types for ENCODE4, CATlas and ABC sources respectively, which were consistent with main CT-FM results (**Supplementary Table 21**). These results demonstrate the benefits of leveraging CTS SNP-annotations from different sources.

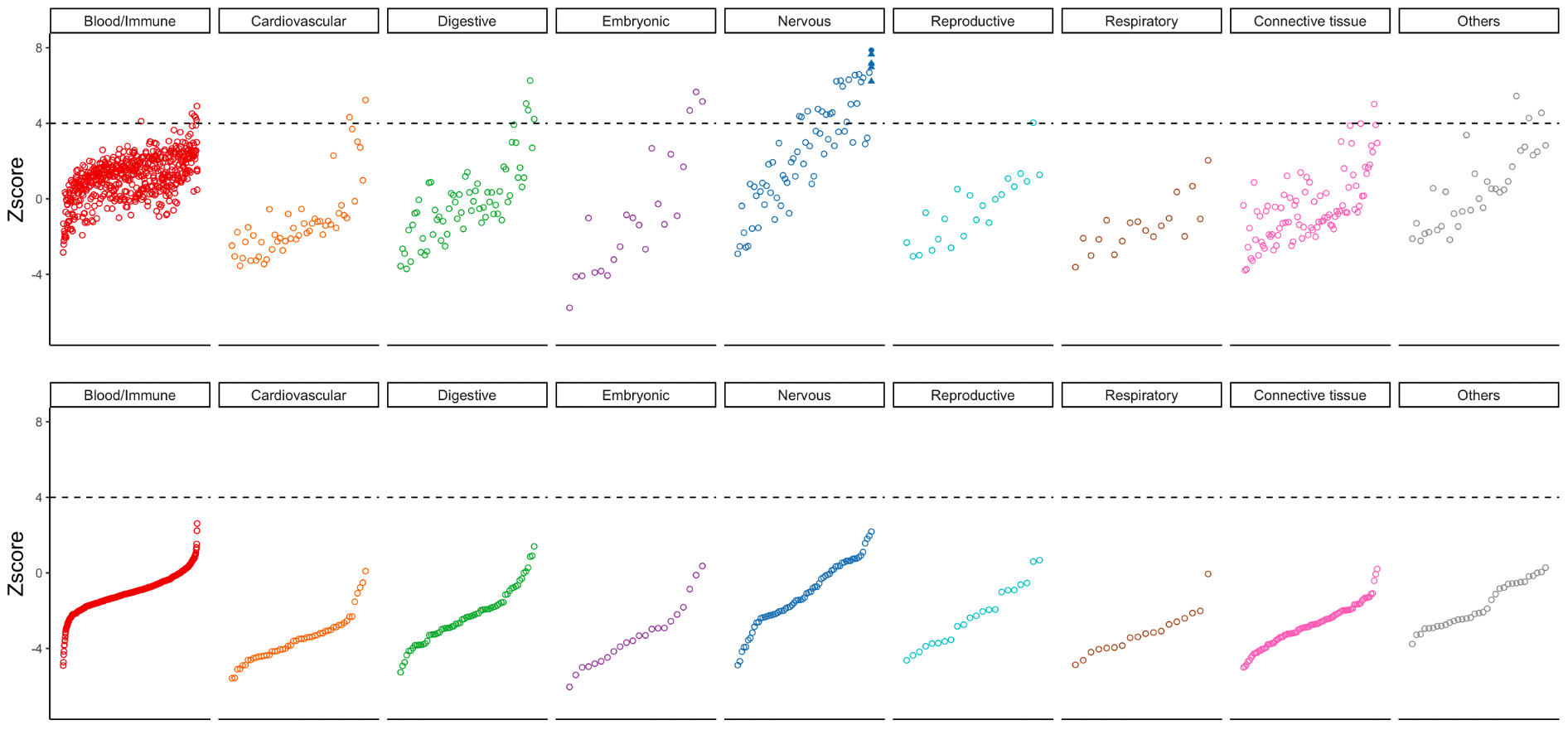
**Supplementary Figure 20. Conditional S-LDSC analysis for BMI.** We report the initial S-LDSC Z-scores of the 924 CTS SNP-annotations used by CT-FM (top). Candidate causal CTS SNP-annotations (i.e., assigned to a CS by CT-FM) are circles (radial glial cells) and triangles (bipolar neurons). To validate that the CSs detected by CT-FM captured most of the conditionally independent causal signal, we reran S-LDSC conditioned on the SNP-annotations in CT-FM CSs (and the background annotations) on the remaining CTS SNP-annotations (i.e., not assigned to a CS by CT-FM). We report the new S-LDSC Z-scores (bottom). We observed that none of the remaining CTS SNP-annotations had a $\tilde{\tau}$ Z-score > 4, confirming that no CTS conditional effect remains in the data after identifying CS with CT-FM.

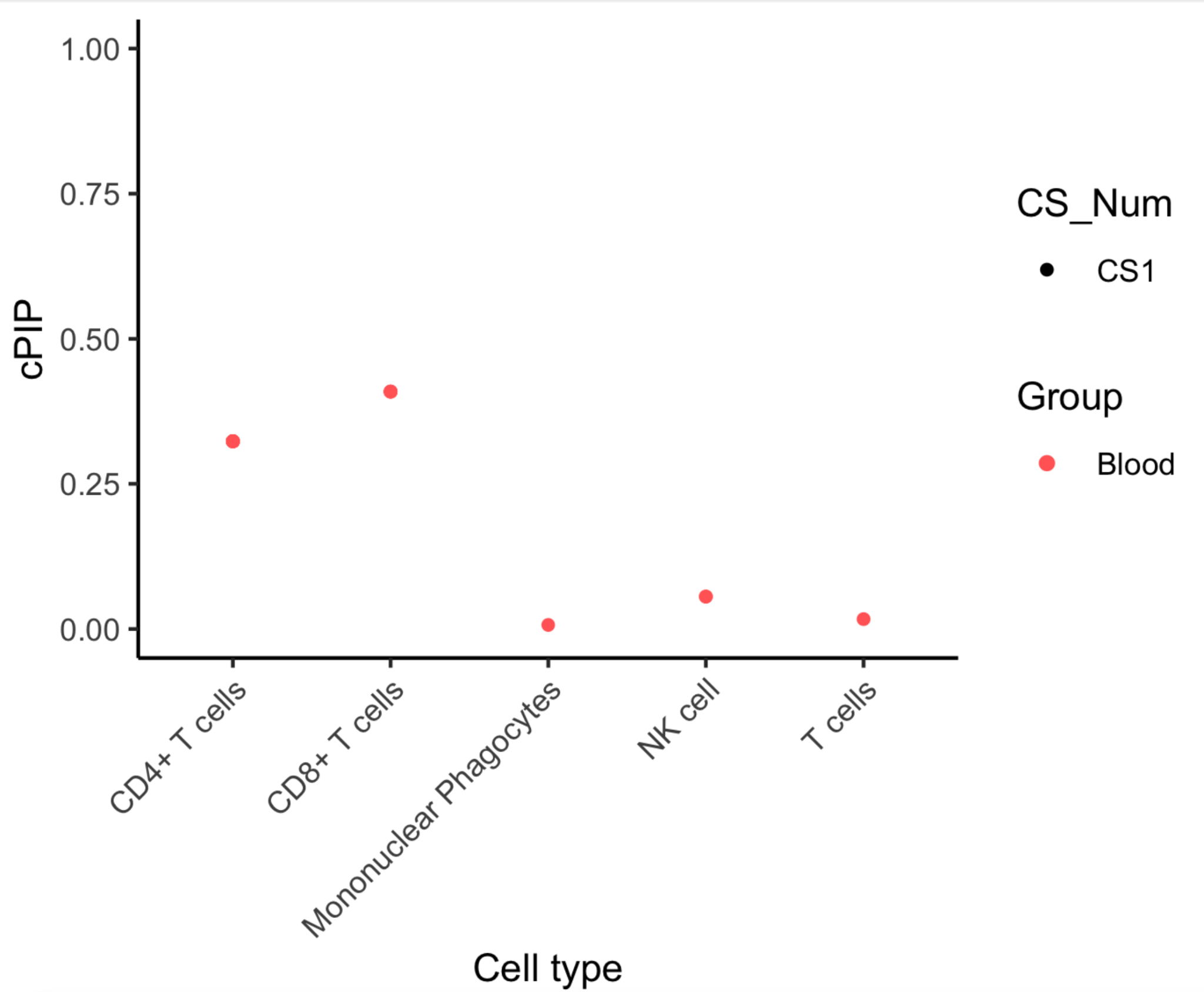

**Supplementary Figure 21. CT-FM results in a second rheumatoid arthritis GWAS** ^23^**.** CT-FM identified one ICS mostly driven by T cells-related CTS SNP-annotations (max PIP = 0.31 for CD8+ T cells). Of note, a second credible set was identified containing B cell-related CTS SNP-annotations (max PIP = 0.27 for naive B cells, data not shown) but was excluded from the analysis during the quality control step (see **Methods**).

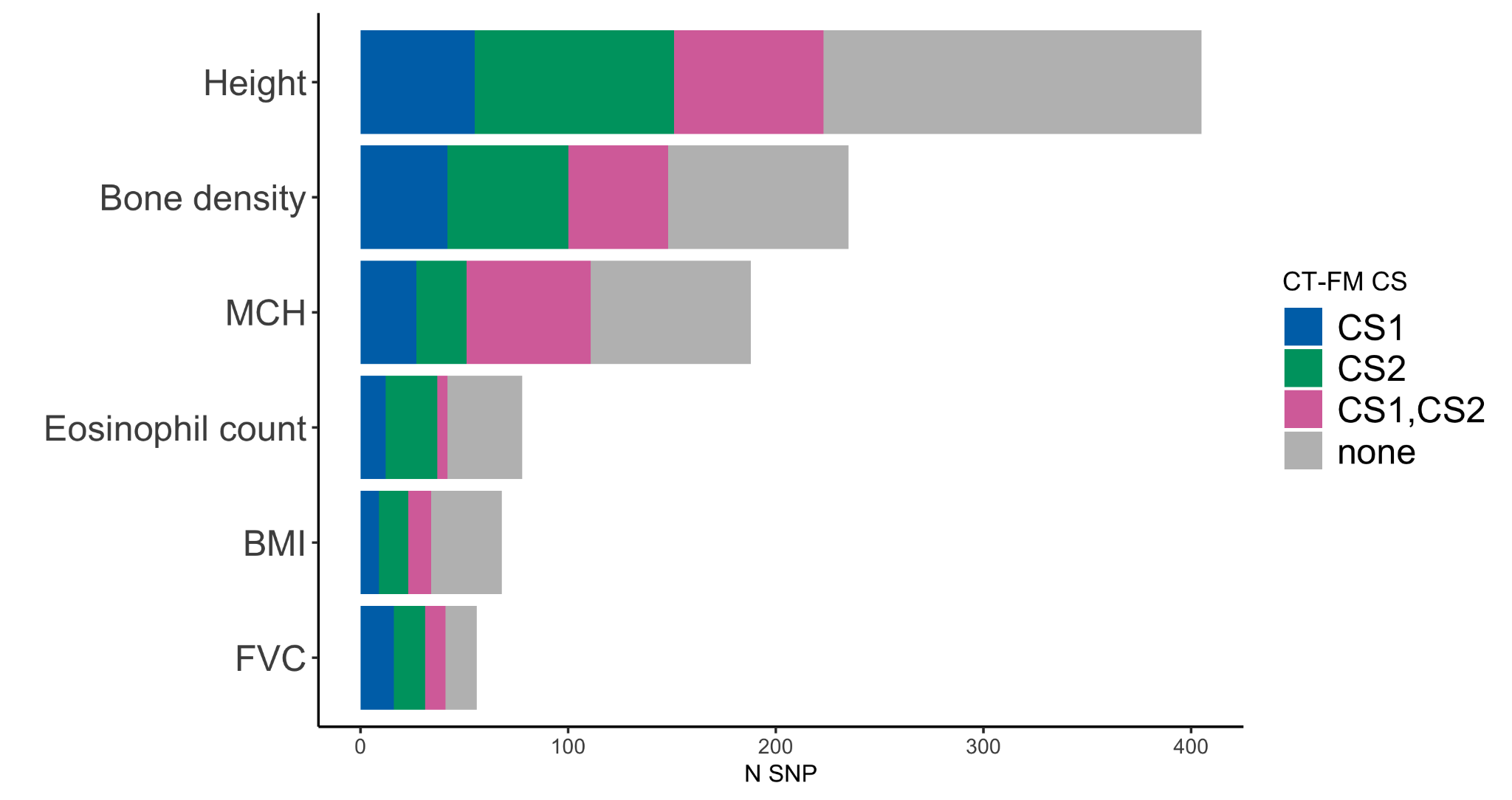

**Supplementary Figure 22. Results of CT-FM-SNP for UK Biobank traits with 2 CT-FM independent causal sets.** For 6 traits with 2 CT-FM credible sets (rows), we indicate the proportion of SNPs assigned to causal CTS SNP-annotations of CT-FM ICS1 (blue), CT-FM ICS2 (green), both CT-FM ICS1 and ICS2 (purple) and the proportion of SNPs assigned to different CTS SNP-annotations not found in CT-FM ICSs. 59% of SNPs were assigned to CTS SNP-annotations previously identified by CT-FM across 6 traits.

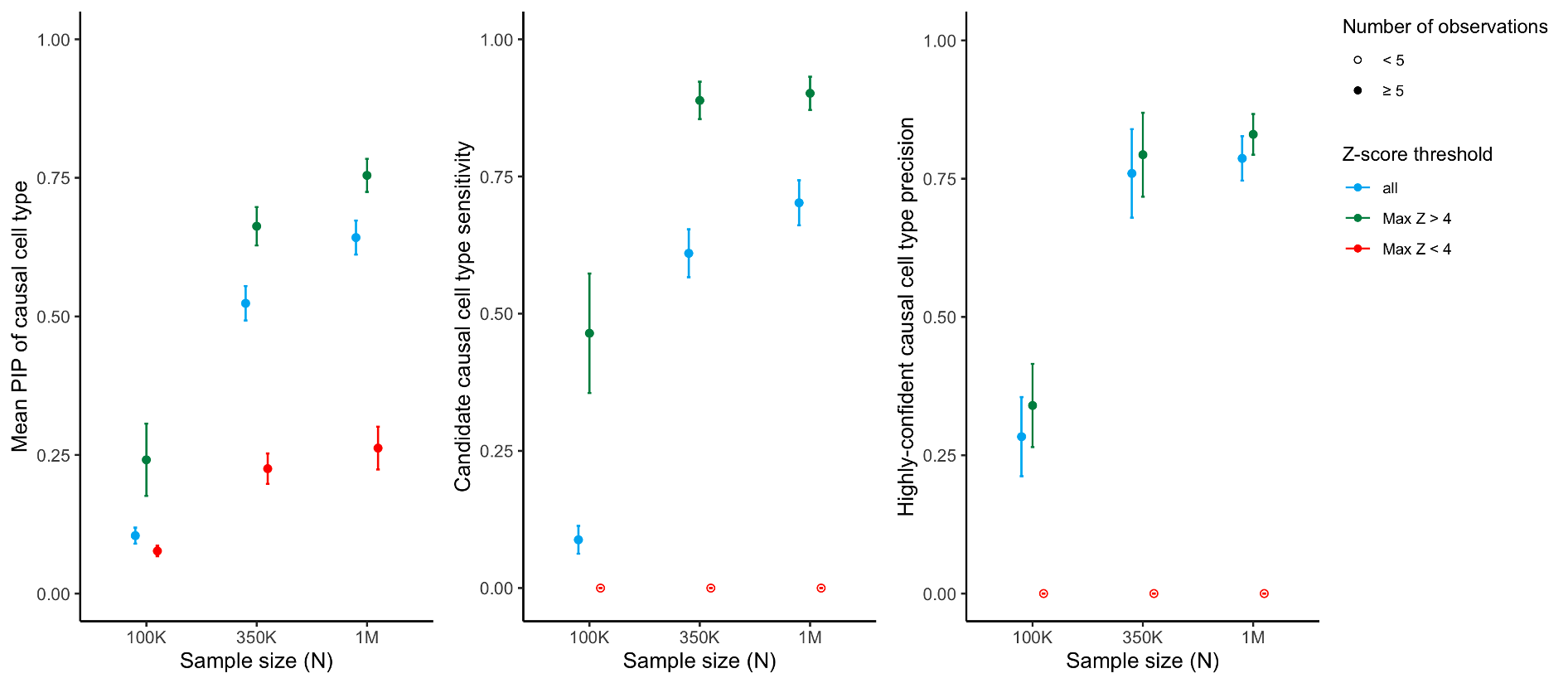
**Supplementary Figure 23. Preliminary simulations to assess CT-FM power and accuracy as a function of S-LDSC maximum Z-score.** We report the power and accuracy of CT-FM in simulations with one causal cell type and different sample sizes, when considering all simulations (blue), simulations where S-LDSC maximum Z-score is > 4 across all the CTS SNP-annotations (green), and simulations where S-LDSC maximum Z-score is ≤ 4 across all the CTS SNP-annotations (red). We report the mean PIP of the causal SNP-annotation(s), the proportion of causal SNP-annotations identified as a candidate causal cell type (candidate causal cell type sensitivity), and the proportion of SNP-annotations identified as a highly-confident causal cell type that are truly causal (highly-confident causal cell type precision). For each scenario, we performed 500 simulations. Error bars represent 95% confidence intervals. We note that these preliminary simulations were performed as described in the Methods section, except that we did not rescaled positive per-SNP h2 after setting negative expected per-SNP h2; indeed, setting those values to 0 led to h2 enrichment lower than observed in the height GWAS, thus leading to many simulations with an S-LDSC maximum Z-score is ≤ 4 (unlike in main simulations) (416, 159, and 114 in simulations with N = 100K, 350K, and 1M, respectively). We observed that restricting CT-FM analyses to simulations where S-LDSC maximum Z-score is > 4 provides fairly high power and accuracy. Because expected per-SNP *h^2^* can be negative, we initially set these values to 0, and rescaled positive per-SNP *h^2^* so that expected *h^2^* enrichment of each annotation in the model was similar to the ones observed on the height GWAS.

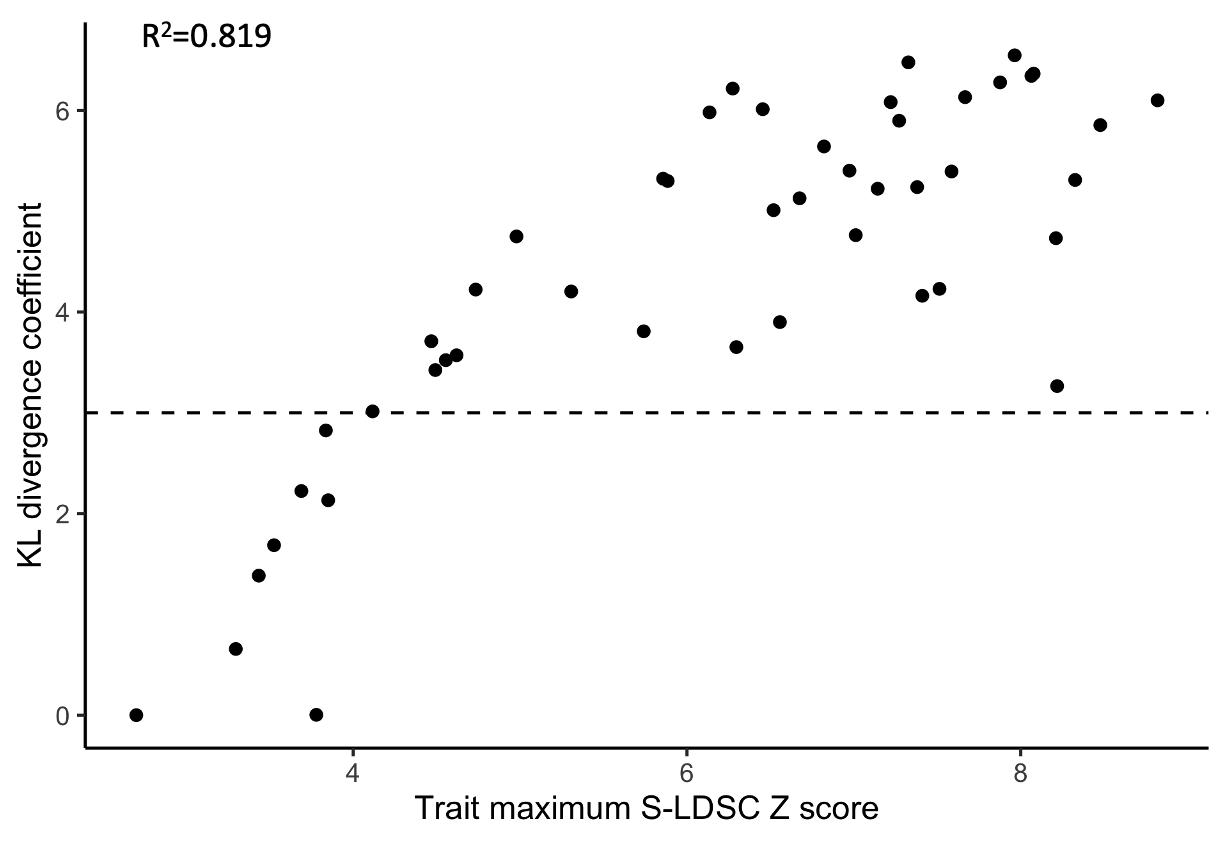

**Supplementary Figure 24. Comparison of KL divergence coefficients with maximum S-LDSC Z-score observed in a SuSiE CS.** We report the results of a permissive CT-FM analysis (no S-LDSC Z-score threshold) for 47 UK Biobank traits. We observed a strong correlation (R2=0.82) between the KL divergence coefficient (y-axis) and the maximum S-LDSC Z-score observed in a SuSiE CS (x-axis). The threshold of KL coefficient ≥ 3 (dashed line) was used in this study as it optimizes the selection of well-powered credible sets.

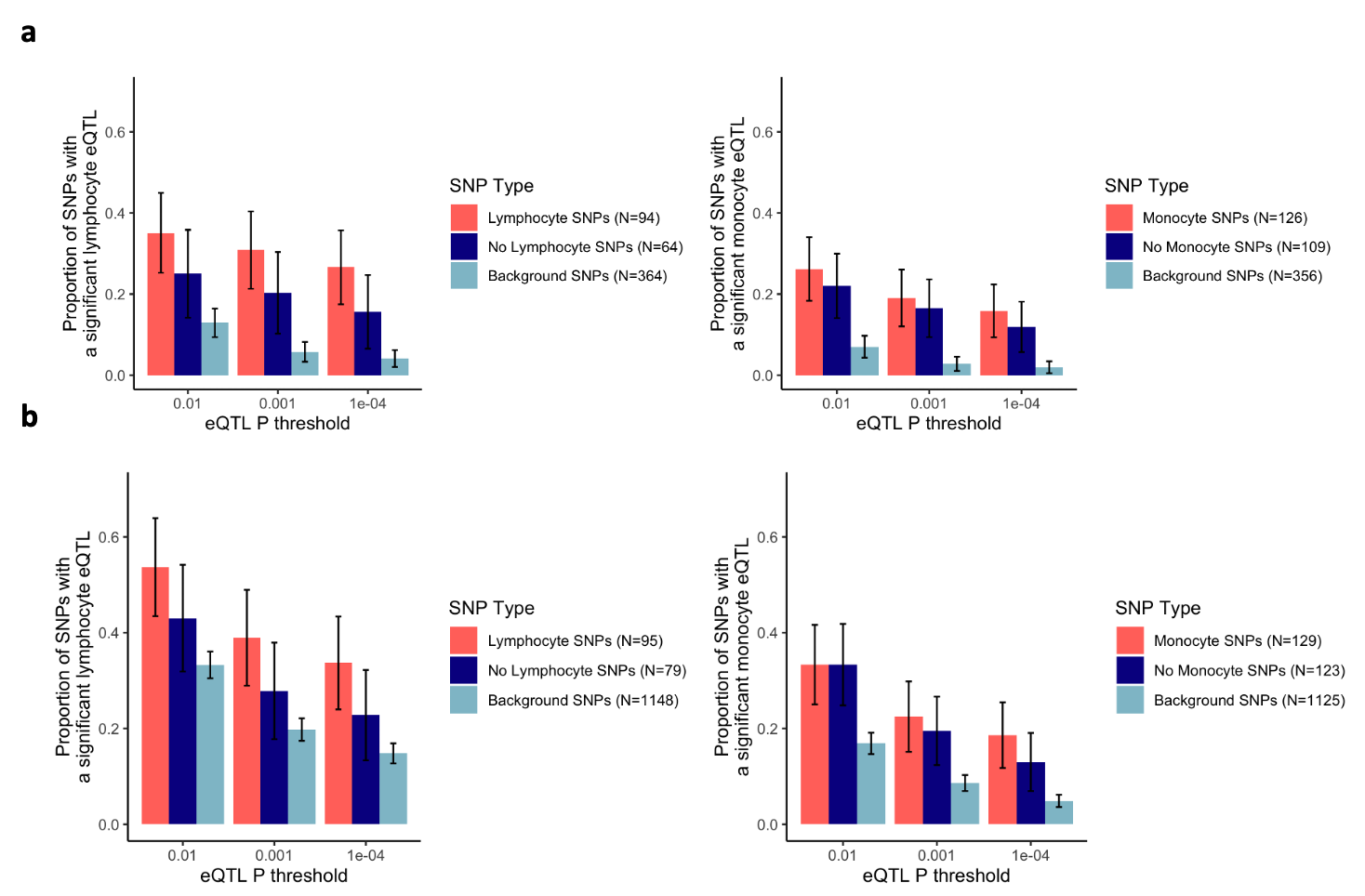

**Supplementary Figure 25. Validating CT-FM-SNP using single-cell cis-eQTLs from OneK1K.** We report the fraction of lymphocyte count candidate SNPs assigned to a lymphocyte cell type by CT-FM-SNP that is a lymphocyte single-cell cis-eQTL in OneK1K and the fraction of monocyte count candidate SNPs assigned to a monocyte cell type by CT-FM-SNP that is a monocyte single-cell cis-eQTL in OneK1K (y axis) at different eQTL p-value thresholds (x axis). We compare these proportions to 1) lymphocyte/monocyte candidate SNPs which were not assigned to a relevant cell type by CT-FM-SNP (no lymphocyte / no monocyte SNPs) and to 2) background SNPs from 16 traits not genetically correlated to lymphocyte count and monocyte count as the baseline. Error bars represent 95% confidence intervals. **(a)** Proportions when restricting eGenes to lymphocyte/monocyte count cS2G genes **(b)** Proportions when restricting eGenes to all protein-coding genes.

25.	Zou, Y. Diagnostic for fine-mapping with summary statistics. <https://stephenslab.github.io/susieR/articles/susierss_diagnostic.html>.
